# Potential strategies for strengthening surveillance of lymphatic filariasis in American Samoa after mass drug administration: targeting older age groups, hotspots, and household members of infected persons

**DOI:** 10.1101/2020.04.07.20056549

**Authors:** Colleen Lau, Meru Sheel, Katie Gass, Saipale Fuimaono, Michael David, Kimberly Won, Sarah Sheridan, Patricia Graves

## Abstract

**Background:** Under the Global Programme to Eliminate Lymphatic Filariasis (LF), American Samoa conducted mass drug administration (MDA) from 2000-2006. Despite passing Transmission Assessment Surveys (TAS) in 2011/2012 and 2015, American Samoa failed TAS-3 in 2016, with antigen (Ag) prevalence of 0.7% (95%CI 0.3-1.8%) in 6-7 year-olds. A 2016 community survey (Ag prevalence 6.2% (95%CI 4.4-8.5%) in age ≥8 years) confirmed resurgence. Here, we explore the potential of targeted strategies to strengthen post-MDA surveillance.

**Methodology/Principal Findings:** Using Ag data plus new antibody data (Wb123, Bm14, Bm33) from the 2016 surveys, we identified risk factors for seropositivity using multivariable logistic regression. We compared TAS with strategies that targeted high-risk subpopulations (older ages, householders of Ag-positive TAS children [index children]) and/or known hotspots, and used NNTest^av^ (average number needed to test to identify one positive) to compare sampling efficiency.

Antibody prevalence in TAS-3 (n=1143) were 1.6% for Bm14 (95%CI 0.9-2.9%), 7.9% for Wb123 (95%CI 6.4-9.6%), and 20.2% for Bm33 (95%CI 16.7-24.3%); and in the community survey (n=2507), 13.9% for Bm14 (95%CI 11.2-17.2%), 27.9% for Wb123 (95%CI 24.6-31.4%), and 47.3% for Bm33 (95%CI 42.1-52.6%). Ag prevalence was 20.7% (95%CI 9.7-53.5%) in households of index children. Higher Ag prevalence was found in males (adjusted odds ratio [aOR] 3.01), age ≥18 years (aOR 2.18), residents of Fagali’i (aOR 15.81), and outdoor workers (aOR 2.61). Using Ag, NNTest^av^ ranged from 142.5 for TAS, to <5 for households of index children. NNTest^av^ was lower in older ages, and highest for Ag, followed by Bm14, Wb123 and Bm33 antibodies.

**Conclusions/Significance:** We propose a multi-stage surveillance strategy, starting with population-representative sampling (e.g. TAS), followed by targeted strategies in subgroups and locations with low NNTest^av^. This approach could potentially improve the efficiency of identifying remaining infected persons and residual hotspots. The utility of antibodies in surveillance should also be explored.

**AUTHOR SUMMARY:** Lymphatic filariasis (LF) is a parasitic infection transmitted by mosquito bites. Globally, tens of millions are infected, with many disfigured and disabled by severe damage to their lymphatic systems, such as severe swelling of the legs (elephantiasis) or scrotum (hydrocele). The Global Programme to Eliminate LF (GPELF) aims to interrupt disease transmission through mass drug administration (MDA), and to control illness and suffering in affected persons. The World Health Organization recommends conducting Transmission Assessment Surveys (TAS) in school children aged 6 to 7 years, to determine if infection rates have dropped to levels where disease transmission is no longer sustainable. From 2000-2006, American Samoa conducted MDA and made significant progress towards eliminating LF. However, despite passing TAS in 2011/2012 and 2015, surveys in 2016 showed evidence of resurgence. Our study explored alternative surveillance strategies and compared their efficiency with TAS. Based on our findings, we recommended that in addition to TAS, strategies that target high-risk populations and hotspots would strengthen surveillance and help countries achieve their goals of LF elimination.

## INTRODUCTION

Lymphatic filariasis (LF) is a mosquito-borne parasitic disease caused by *Wuchereria bancrofti* and *Brugia* species. The Global Programme to Eliminate Lymphatic Filariasis (GPELF), established by the World Health Organization (WHO) in 2000, aims to eliminate the disease as a public health problem through annual mass drug administration (MDA) and alleviation of morbidity and disability in affected persons. GPELF has made enormous progress; by 2017, the program had delivered 7.1 billion drug treatments to >890 million people in 67 of the 72 LF-endemic countries around the world (1). By 2019, 11 countries have been validated by WHO as having eliminated LF as a public health problem (1). To determine whether transmission has reached sufficiently low levels so that MDA can be safely stopped, WHO currently recommends using Transmission Assessment Surveys (TAS), which use critical cut-off numbers of antigen (Ag) positive children aged 6-7 years to determine if transmission has been interrupted in defined evaluation units (2). After passing the first TAS, repeated surveys are recommended every 2-3 years for ongoing post-MDA surveillance.

Although TAS has been a useful tool for making decisions to stop MDA, and for post-MDA surveillance, there is emerging evidence that the current TAS guidelines may not be sufficiently sensitive for identifying residual foci or hotspots of ongoing transmission, especially in the post-MDA setting when prevalence has reached very low levels and the geographic distribution of residual infectious is highly heterogenous (e.g. localised hotspots where infection prevalence is significantly higher than the rest of the surveillance area). This concern has been reported by studies from diverse settings around the world including American Samoa (3-6), Samoa (7), Sri Lanka (8), India (9), and Haiti (10, 11), suggesting that additional and/or alternative surveillance strategies are required to identify residual transmission hotspots, and also provide earlier signals of any resurgence so that programmatic gains can be protected, and long-term success of the GPELF ensured.

The GPELF is one of the largest and most ambitious public health intervention programs in the world, and it is inevitable that new and unexpected challenges will emerge as countries progress through the program. As the GPELF has developed and evolved over the years, operational research has provided valuable evidence to inform and improve its many activities, including diagnostics, surveillance, monitoring, and evaluation (12). Currently, the WHO and GPELF have identified standardised cost-effective surveillance strategies as one of the most important operational research needs (13). Because population-based surveys are resource intensive, potential ways of improving their cost-effectiveness include integrating post-MDA surveillance with other public health programs, and opportunistic screening of populations, e.g. during blood donation, antenatal visits, and pre-employment tests. Other approaches include more sensitive surveillance strategies, such as the detection of filarial antibodies (which may provide earlier indication of transmission and recrudescence compared to Ag) (14, 15), and molecular xenomonitoring (detection of filarial DNA in mosquitoes) (16-19).

American Samoa, a US territory in the South Pacific, is a group of small remote islands where LF is endemic. Infection is caused by *W. bancrofti* and transmitted predominantly by the highly efficient day-biting *Aedes polynesiensis*, as well as other day- and night-biting *Aedes* species. As part of the Pacific Programme to Eliminate LF (PacELF), a baseline survey in 1999 found an Ag prevalence of 16.5% in 18 villages in American Samoa, as measured by the Alere rapid immunochromatographic test (ICT) (20). Annual rounds of MDA were distributed from 2000-2006 (6, 21), and American Samoa passed TAS-1 and TAS-2 in 2011/2012 and 2015, respectively. Two Ag-positive children were identified in TAS-1, and one identified in TAS-2; the critical cut-off for passing TAS for both surveys was a maximum of six children (15). According to current guidelines, passing TAS provides an indication that prevalence has reduced to a level where transmission is thought to be no longer sustainable. However, despite passing both TAS-1 and TAS-2, multiple research studies conducted in American Samoa during the same time period raised suspicions of ongoing transmission. The studies included opportunistic screening of a serum bank collected from adults who were representative of the general population (3), testing adults at a work place and a pre-employment clinic (4, 22), testing residents in suspected hotspots (4), and molecular xenomonitoring of mosquitoes (23). Clustering and persistence of antibody responses were noted in schools in TAS-1 and TAS-2, and may also have been an indication of ongoing transmission (15).

In 2016, American Samoa failed TAS-3 (5), confirming that the earlier suspicions of ongoing transmission were indeed correct, and that in hindsight, resurgence could potentially have been detected earlier using surveillance strategies other than or in addition to the standard TAS. In parallel with TAS-3 in 2016, a community-based survey of 2507 people aged 8 years and older in randomly selected villages found an estimated overall Ag prevalence of 6.2% (95% CI 4.5-8.6%), further confirming resurgence (5). The 2016 studies found that the community-based survey was logistically more difficult and more expensive than the school-based survey, but provided more detailed epidemiological information, including locations of transmission hotspots. Comparison of the results of the parallel surveys found that although the school-based survey had limited sensitivity and negative predictive value for identifying villages with ongoing transmission, specificity and positive predictive value were high (5). In other words, although TAS may not be sufficiently sensitive for detecting all transmission hotspots, the residential locations of Ag-positive school children could lead us to at least some of the reservoirs of ongoing transmission, where more targeted and/or intensive surveillance could potentially be valuable.

According to current WHO guidelines for TAS, for any given population size in an evaluation unit, both the sample size and the threshold number for “passing” are fixed. Therefore, sensitivity and cost-effectiveness of TAS for detecting residual infections and finding potential hotspots will gradually erode over time as prevalence decreases. In other words, if population representative sampling is used for surveillance (e.g. the current TAS protocols), the average number of people who need to be tested in order to find one abnormal result, or ‘Number Needed to Test’ (NNTest^av^) will increase as infection prevalence decreases. The NNTest is similar in concept to the commonly used ‘Number needed to screen’ (NNS) and ‘Number needed to treat’ (NNT) measures (24, 25). The advantages of NNS and NNT measures over more traditional metrics such as relative risk, relative risk reduction, and odds ratios is that they are more easily understood by clinicians and patients, and intuitively more useful for decision making. While NNS and NNT are typically used to measure the benefit of screening or treatment on a specific clinical outcome, we use NNTest^av^ here only as a measure of the probability of identifying a positive test. In this context, NNTest^av^ could be used to measure and rank the relative sensitivity and efficiency of different surveillance strategies or combinations of strategies, e.g. population representative sampling vs strategies that target high-risk populations and/or locations, using different serological markers, and in different age groups. NNTest^av^ could be used as a summary measure to compare the conditional probabilities of identifying a positive test in subgroups, e.g. positive antigen in 6-7 year-old children who live in known LF hotspots vs positive antibody in adults in randomly selected villages.

It is important to note that strategies that minimise NNTest^av^ may not provide the data required to accurately estimate population-level infection prevalence, but are more likely to identify rare events (if they exist) such as Ag-positive people in the post-MDA setting. For post-MDA surveillance, identifying any remaining Ag-positive people (which might provide indications of hotspots and/or resurgence) is arguably strategically more important than having accurate population-level estimates of infection prevalence. In American Samoa, considering that LF prevalence is disproportionately higher in some groups (e.g. males, older individuals) (3) and transmission has been shown to be highly focal in the post-MDA setting (3-5, 7, 8), targeted surveillance of high-risk persons and locations could significantly reduce the NNTest, thereby improving the precision and cost-effectiveness of post-MDA surveillance.

In this paper, we use American Samoa as a case study to explore potential options for reducing NNTest for post-MDA surveillance. We focus on strategies that start with population representative sampling (e.g. surveying school-aged children through TAS) and using the findings to inform further targeted surveillance of household and community members and/or surveillance of older school aged children. Using school-based surveys for the initial phase of screening enables large populations to be tested at relatively low cost and is generally logistically simpler than large community-based surveys. Furthermore, TAS is already a WHO-approved tool that country programs are familiar with and have the resources to implement. Current TAS guidelines recommend that all Ag-positive cases should be treated and tested for microfilaraemia (2). However, there are no evidence-based recommendations for using information about Ag-positive children from the TAS to inform further surveillance activities. Considering that young children spend most of their time at home or at school, along with our previous findings that LF transmission hotspots are highly focal in American Samoa (3-5), we hypothesize that Ag-positive children identified through TAS (index children) could be used as sentinels for more targeted and therefore more cost-effective surveillance.

This study aims to identify risk factors for LF infection in American Samoa and use this information to compare NNTest for different surveillance strategies. The specific objectives of the study are to determine if, compared to standard TAS of 6-7 year-old children, NNTest can be reduced by targeted surveillance of subpopulations with higher infection prevalence such as older age groups, household members of Ag-positive children identified in TAS, residents of known transmission hotspots, or a combination of the above strategies.

## METHODS

### A. Study location

American Samoa consists of a group of small remote islands in the South Pacific, with a total population of ∼55,519 (26). Over 95% of the population live on the largest island of Tutuila and the adjacent island of Aunu’u. Very small populations reside in the remote Manu’a islands, which were not included in this study.

### B. Informed consent, ethics approvals, and cultural considerations

Signed informed consent was obtained from adult participants or from parents/ guardians of those aged <18 years, along with verbal assent from minors. The study was conducted in collaboration with the American Samoa Department of Health, and official permissions for school and village visits were granted by the Department of Education and the Department of Samoa Affairs, respectively. All field activities were carried out in a culturally appropriate and sensitive manner with bilingual local field teams, and with verbal approval from village chiefs or mayors prior to conducting the community surveys. Surveys were conducted in English or Samoan depending on the participants’ preference. Ethics approvals were granted by American Samoa Institutional Review Board, and the Human Research Ethics Committee at the Australian National University (protocol number 2016/482). The Institutional Review Board of the U.S. Centers for Disease Control and Prevention (CDC) determined CDC to be a non-engaged research partner.

### C. Study components

The 2016 field study included four components, as summarised in Figures 1 and 2.

**Figure 1.**
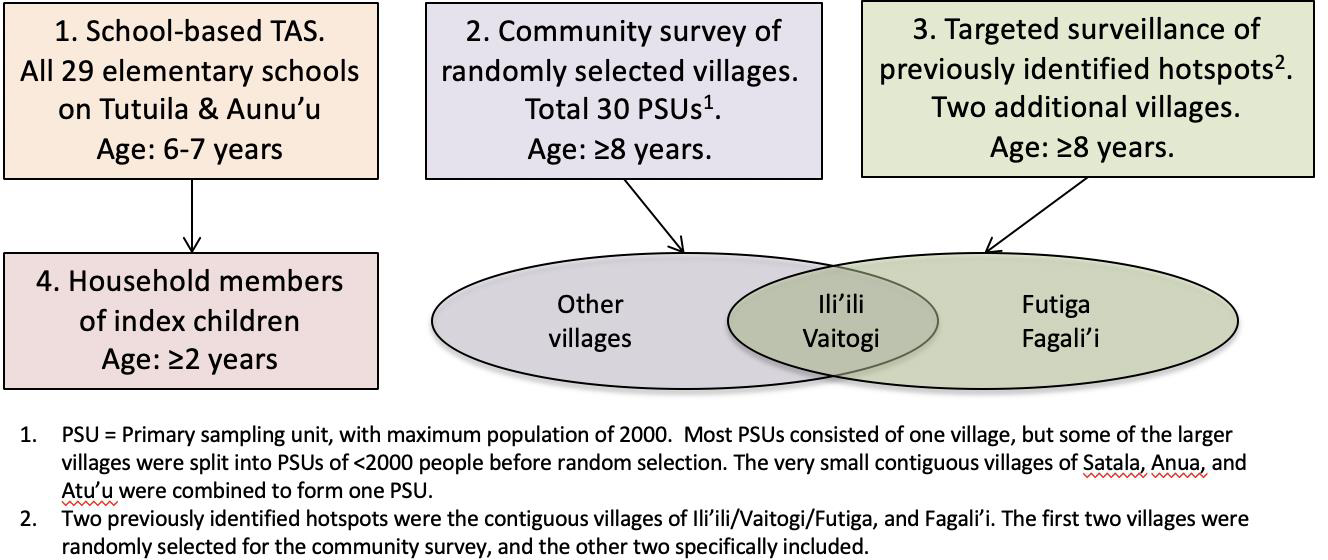
Summary of study components, American Samoa 2016.

**Figure 2.**
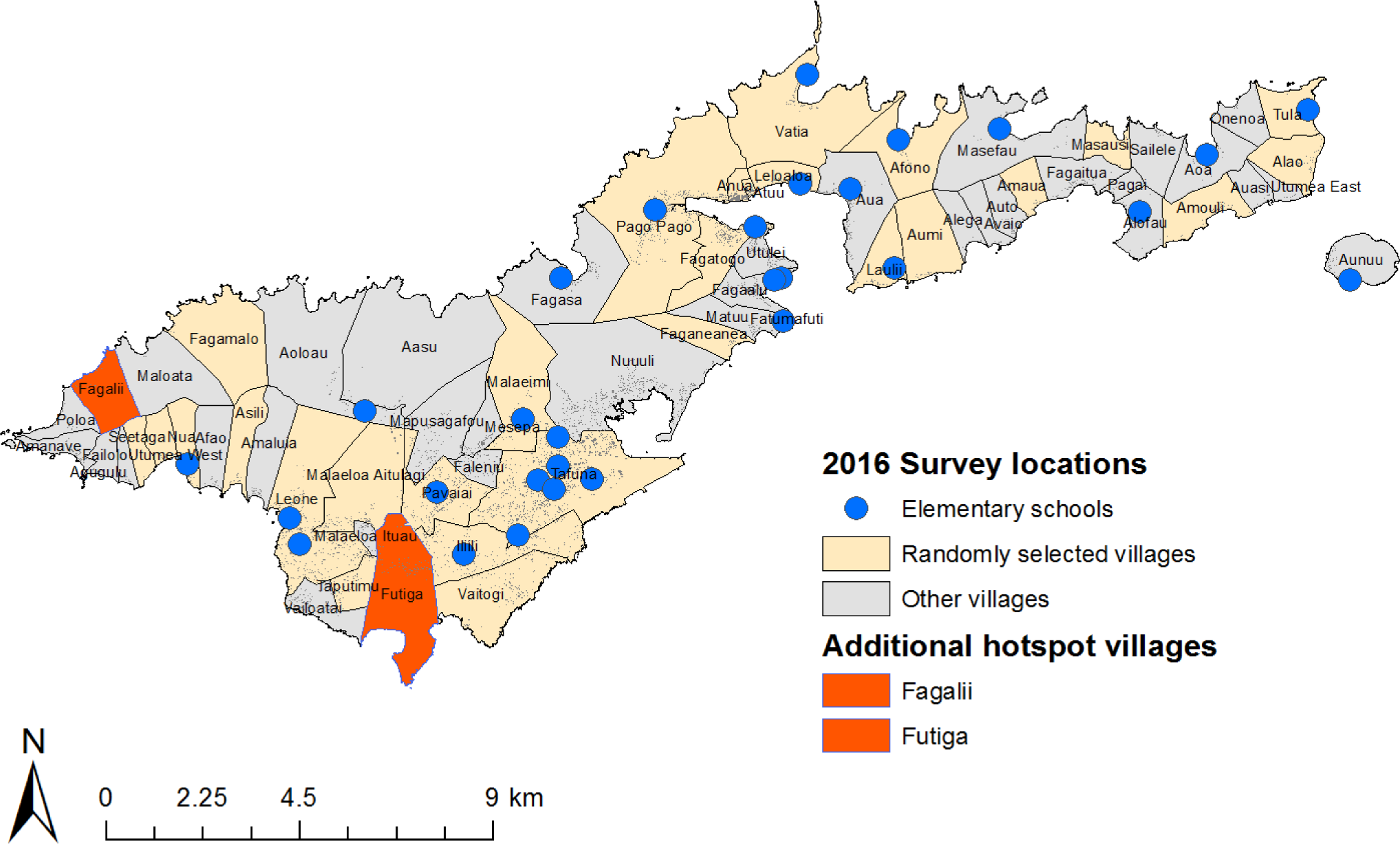
Survey locations in American Samoa 2016: elementary schools (blue circles), randomly selected villages (yellow), and additional hotspot villages (orange).

1. School-based TAS-3 of 6-7 year-old children
2. Community survey of randomly selected villages
3. Targeted surveillance in two previously identified hotspots (total of 4 villages, two of which were also randomly selected for component 2 above)
4. Household members of Ag-positive children identified in TAS (index children)

Descriptions of each component are provided below and details on sampling design and results of components 1 and 2 have been previously published (5).

#### C1. School-based TAS-3 of 6-7 year-old children

In 2016, we conducted a school based TAS in American Samoa according to WHO guidelines. As previously described (5), the survey included Grade 1 and 2 children from all 29 elementary schools on the islands of Tutuila and Aunu’u (Figure 2). Based on WHO guidelines, the critical cut-off for passing TAS-3 was a maximum of six Ag-positive children (2).

#### C2. Community-based survey of randomly selected villages (age ≥8 years)

In parallel with TAS-3, a community-based population proportionate survey was conducted in 30 randomly selected primary sampling units (PSUs) (Figure 2) (5). Most PSUs consisted of one village, but some of the larger villages were split into PSUs of <2000 people, and the very small contiguous villages of Satala, Anua, and Atu’u were combined to form one PSU. Households were randomly selected within each village, and all household members aged ≥8 years were invited to participate.

#### C3. Targeted surveillance of previously identified hotspots

Our previous studies identified (3) and later confirmed (4) two transmission hotspots in American Samoa, located in the contiguous large villages of Ili’ili, Vaitogi and Futiga, and the small remote village of Fagali’i (Figure 2). The two Ag-positive children identified in TAS-1 and the one Ag-positive child identified in TAS-2 all attend the same elementary school located in the first hotspot. The villages of Ili’ili and Vaitogi were randomly selected for the 2016 community-based survey described above, and Futiga and Fagali’i were specifically added to the survey to allow further assessment of the hotspots. In the two additional villages, the survey was conducted using the same methods as the randomly selected villages. In Fagali’i, participants also included volunteers from households that were not randomly selected; demographics and infection rates did not differ significantly between volunteers and randomly selected community members, and the two groups were combined for estimates of village-level seroprevalence.

#### C4. Household and community members of Ag-positive school children from TAS-3

All index children from TAS-3 were followed up at home and provided with treatment with diethylcarbamazine (DEC) and albendazole according to WHO guidelines (see below). All household members of index children who were aged ≥2 years and present during the home visits were invited to participate in the study, using the same survey methods as the community-based survey. If households had already been sampled as part of the randomly selected households for the community survey, household members aged 2-7 years and others who had not already been tested during the community survey were also invited to participate.

### D. Questionnaires and household locations

Standardised electronic questionnaires were used to collect demographic and household data, including gender, age, work location, time lived in American Samoa, ever taken MDA, travel in past 12 months, and living in a known hotspot. Work location was classified as indoor, outdoor, tuna cannery (largest private employer in American Samoa), and other (including mixed indoor/outdoor, unemployed, retired or unknown). For the community survey, hotspots survey, and household visits of index children, GPS locations of households were recorded. Data were collected by bilingual field research assistants (in Samoan or English according to each participant’s preference), using smartphones and the LINKS electronic database system (27).

### E. Blood collection and laboratory tests

The Alere Filariasis Test Strip (FTS) was used to detect circulating filarial antigen. For each participant, a 200uL finger-prick blood sample was collected into heparinised microtainers (BD, North Ryde, NSW Australia), and samples were kept cool and transported to a laboratory where FTS were conducted on the same or next day. For household members of index children, samples were tested during the household visits so that Ag-positive persons could be provided with treatment at that time, and avoid the need for additional visits. For Ag-positive persons, microfilaria (Mf) slides were prepared using standard procedures (2). Dried blood spots (DBS) were prepared using TropBio filter papers (Cellabs, Sydney, Australia), and shipped to the US CDC for assays of Wb123, Bm14, and Bm33 antibodies using previously described methods (15).

### F. Treatment of antigen-positive persons

Unless medically contraindicated, Ag-positive individuals were offered treatment with DEC (6mg/kg) and Albendazole (400mg). As described above, Ag-positive children identified through TAS-3 were treated at home, in the presence of a parent or guardian. Ag-positive persons identified through the community survey were contacted by phone and invited (in some cases with family members) to attend a clinic for treatment. All medications were provided free of charge.

### G. Definition of subgroups

The following definitions were used for the different subgroups included in the analyses:

- Index child – Ag-positive child identified through TAS
- Index households – household members of index child
- Index villages/communities – villages/communities where at least one index child lived
- Randomly selected households – households selected for the community survey

Figure 3 provides a schematic representation of the definitions of index households, community members of index children, and index communities.

**Figure 3.**
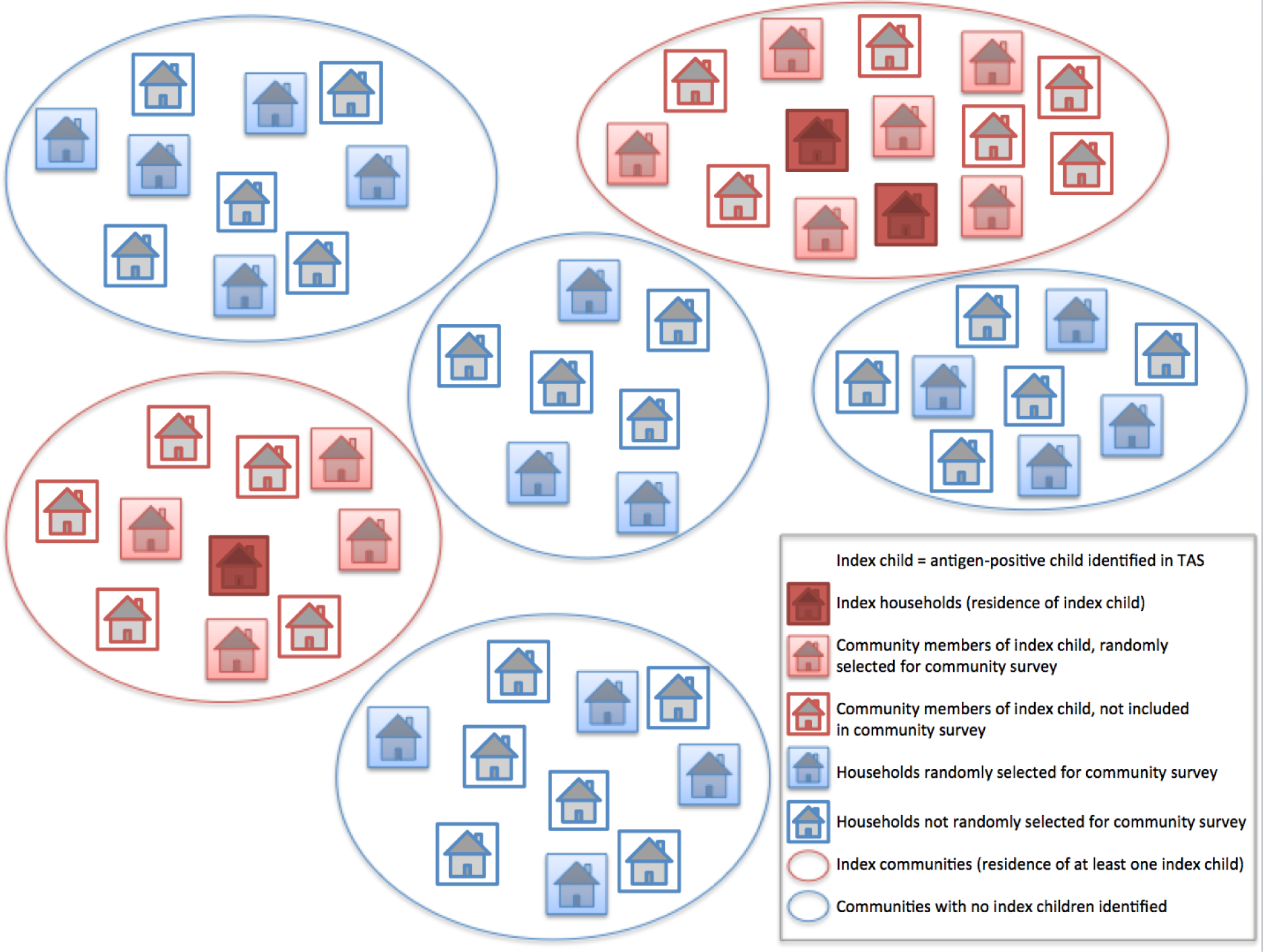
Definitions of subgroups used for analyses

### H. Statistical analysis and mapping

Analyses were performed using Microsoft Excel or Stata version 14 (Stata Corp, College Station, TX), and *p* values of <0.05 were considered statistically significant. Spatial data on island and village boundaries in American Samoa were obtained from the American Samoa Coastal and Marine Spatial Planning Data Portal (28). Geographic information systems (GIS) software (ArcGIS v10.4.1, Environmental Systems Research Institute, Redlands CA) was used to manage spatial data and produce maps.

#### H1. Crude and Age-adjusted Seroprevalence

Crude prevalences were estimated for Ag, Mf, and antibodies (Bm14, Wb123, and Bm33), and 95% confidence intervals (CI) were calculated using the Clopper-Pearson binomial exact method (29). Adjusted seroprevalence for Ag and antibodies (for age, sex, and survey design) were calculated using previously described methods (5). Detailed explanation of the methods used to calculate adjusted seroprevalence are included in S1 Appendix.

Population representative estimates of seroprevalence for 6-7 year-olds were calculated using TAS-3 results, and for the older age groups (≥8 years) using data from the community survey of randomly selected villages. Adjusted seroprevalence of Ag and antibodies were also calculated for the following subgroups based on age, residence in known transmission hotspots, and residential proximity to an index child:

1. Age groups, based on ages that could potentially be targeted by different surveillance strategies:
  - 6-7 year-old children (e.g. routine TAS)
  - 8-12 year-old children (e.g. older elementary school)
  - 13-17 year-old children (e.g. high school)
  - ≥8 years old (e.g. the 2016 community-based survey in American Samoa)
  - ≥18 years old (e.g. surveillance of adults at a work place)
2. Residents of previously identified hotspots, stratified by age groups:
  - Residents of Ili’ili, Vaitogi, Futiga (total population 5877)
  - Residents of Fagali’i (total population 247)
3. Household members of index children (limited to those aged ≥8 years to allow comparison with results from population representative community survey).

These three factors were specifically chosen for exploring the NNTest concept because targeted strategies could be relatively easily developed for these subgroups. In comparison, it would be logistically more difficult to develop surveillance strategies that target other risk factors such as number of years lived in American Samoa, travel history, or previous participation in MDA.

#### H2. Risk factor analysis and logistic regression models

We undertook descriptive analyses of demographic and behavioural data collected through questionnaires, and determined statistical differences between proportions using Pearson’s chi squared tests, Fisher’s exact tests, or two-sample test of proportions (Stata command prtesti). Logistic regression was used to assess associations between questionnaire variables and Ag, Mf, and each antibody. Any variables with p <0.2 on univariate analyses were tested using multivariable logistic regression. Variables with p ≥0.2 were sequentially removed from the multivariable models to arrive at the most parsimonious models, and variables with p <0.05 were retained in the final model.

#### H3. Intra-class correlation

Intra-class correlation (ICC) was used to provide a measure of the degree of clustering for Ag and each Ab within PSUs and households. ICCs were estimated using a multi-level logistic regression model (Stata commands ‘melogit’ and ‘estat icc’) with age and sex included as fixed effects, and three levels: PSUs, households, and household members. ICC values range from zero to one and provides a measure of how similar observations are at each level, in this case PSUs and households. A higher ICC indicates that the outcome measure is more homogenous (i.e. high degree of clustering) at that level.

#### H4. Number needed to test

For Ag and each antibody, three NNTest measures were calculated using the following formula (where p = adjusted seroprevalence for each subgroup):

p = adjusted seroprevalence, N = sample size (NNTest), and probability of detecting at least one case = 1-(1-p)^N

- NNTest^av^. Average number needed to test in order to identify one positive result: 1/p.
- NNTest^50^. Number (N) needed to test to provide a 50% chance of identifying at least one positive result, where 1-(1-p)^N = 0.5, i.e. NNTest^50^ = *ln* (0.5) / *ln* (1-p)
- NNTest^95^. Number (N) needed to test to provide a 95% chance of identifying at least one positive result, where 1-(1-p)^N = 0.95, i.e. NNTest^95^ = *ln* (0.05) / *ln* (1-p)

NNTest^av^ was not calculated for Mf because it is impractical to use Mf as a screening tool. It is important to note that NNTest^av^ itself does not provide an indication of the sample size required for surveys, and if NNTest^av^ is *n* for a specific strategy, testing *n* number of people certainly would not guarantee that an infected person will be identified. NNTest^50^ and NNTest^95^ are used here as examples of the sample size required to providing different degrees of certainty (50% and 95% certain) that positive cases are not missed.

## RESULTS

### A. Seroprevalence of antigen and antibodies

#### A1. School-based TAS-3 of 6-7 year-old children

Antigen results from the school-based TAS-3 and the community survey of randomly selected villages have been previously described in detail (5). Briefly, of the 1143 school children tested in TAS-3, 51.2% were female, and 90.4% were aged 6-7 years old. The children represented 52.4% of all Grade 1 and 2 enrolments and were highly representative of 6-7 year-olds in American Samoa. Nine Ag-positive children (index children) were identified, and one of them was Mf-positive. After adjusting for sex and participation rates by school, the estimated overall Ag prevalence in American Samoa was 0.7% (95% CI 0.3-1.8%) (5). The nine index children attended five different schools and lived in six different villages (Figure 4). Overall antibody prevalence in TAS-3 were 1.6% for Bm14 (95% CI 0.9-2.9%), 7.9% for Wb123 (95% CI 6.4-9.6%), and 20.2% for Bm33 (95%CI 16.7-24.3%). Five of the nine Ag-positive children (55.6%) were seropositive for all three antibodies, while one child (11.1%) was seronegative for all antibodies.

**Figure 4.**
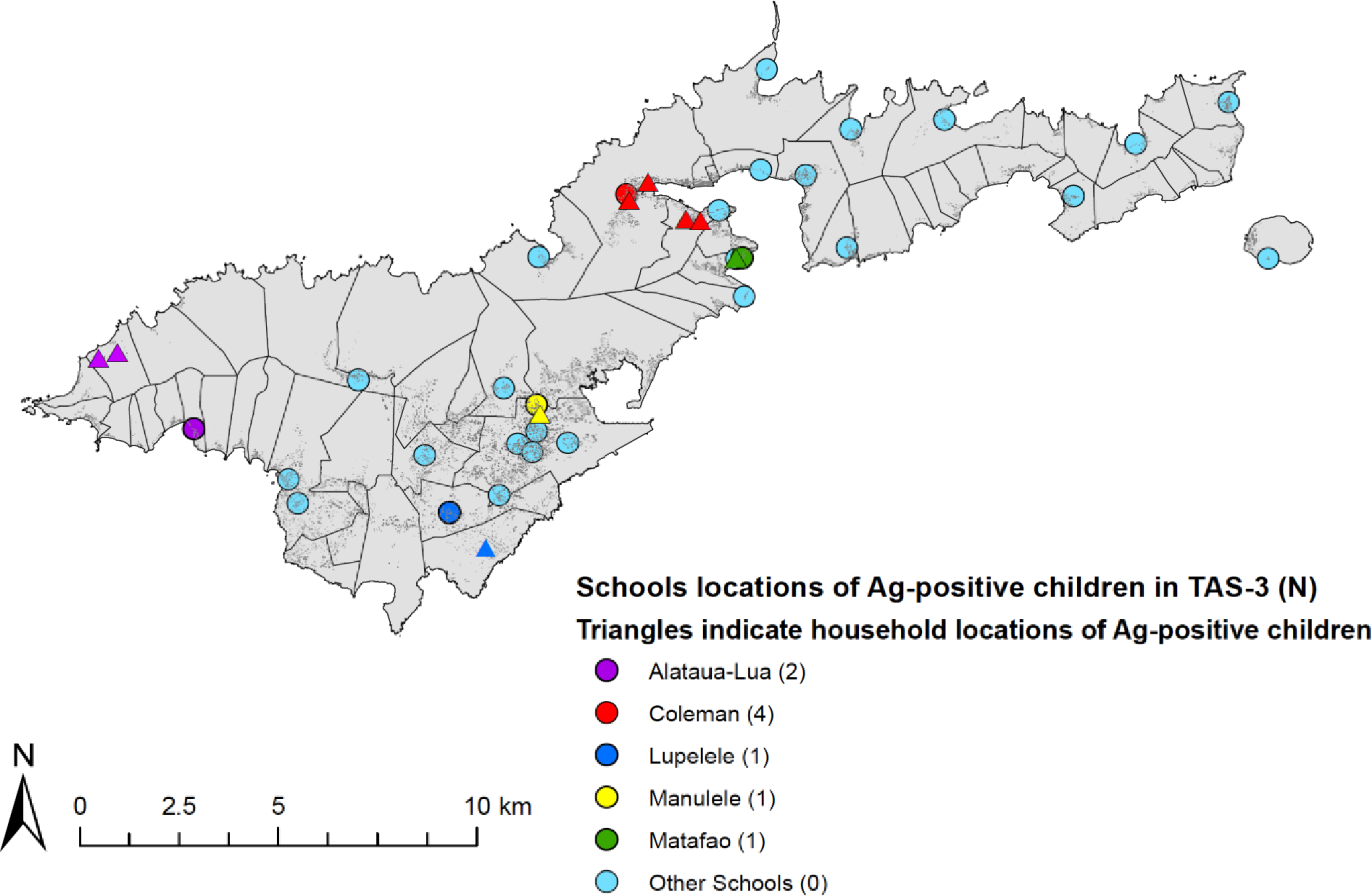
Locations of schools (circles) and households (triangles) of Ag-positive children identified in TAS-3, American Samoa 2016. Colours of triangles indicate the school location of each child.

#### A2. Community-based survey of randomly selected villages (age ≥8 years)

The community survey of 30 randomly selected PSUs included 2507 participants aged ≥8 years, and found a total of 102 Ag-positive persons (5). Of 86 antigen-positive community members where Mf slides were available, 22 (25.6%) were Mf-positive. After adjusting for age, sex, and the survey design, the estimated overall Ag prevalence was 6.2% (95% CI 4.4-8.5%), with village-level Ag prevalence ranging from 0% to 47% (Figure 5). Overall antibody prevalence in the community survey were 13.9% for Bm14 (95% CI 11.2-17.2%), 27.9% for Wb123 (95% CI 24.6-31.4%), and 47.3% for Bm33 (95% CI 42.1-52.6%). The seroprevalence of Ag and antibodies varied significantly between age groups (Figure 6a).

**Figure 5.**
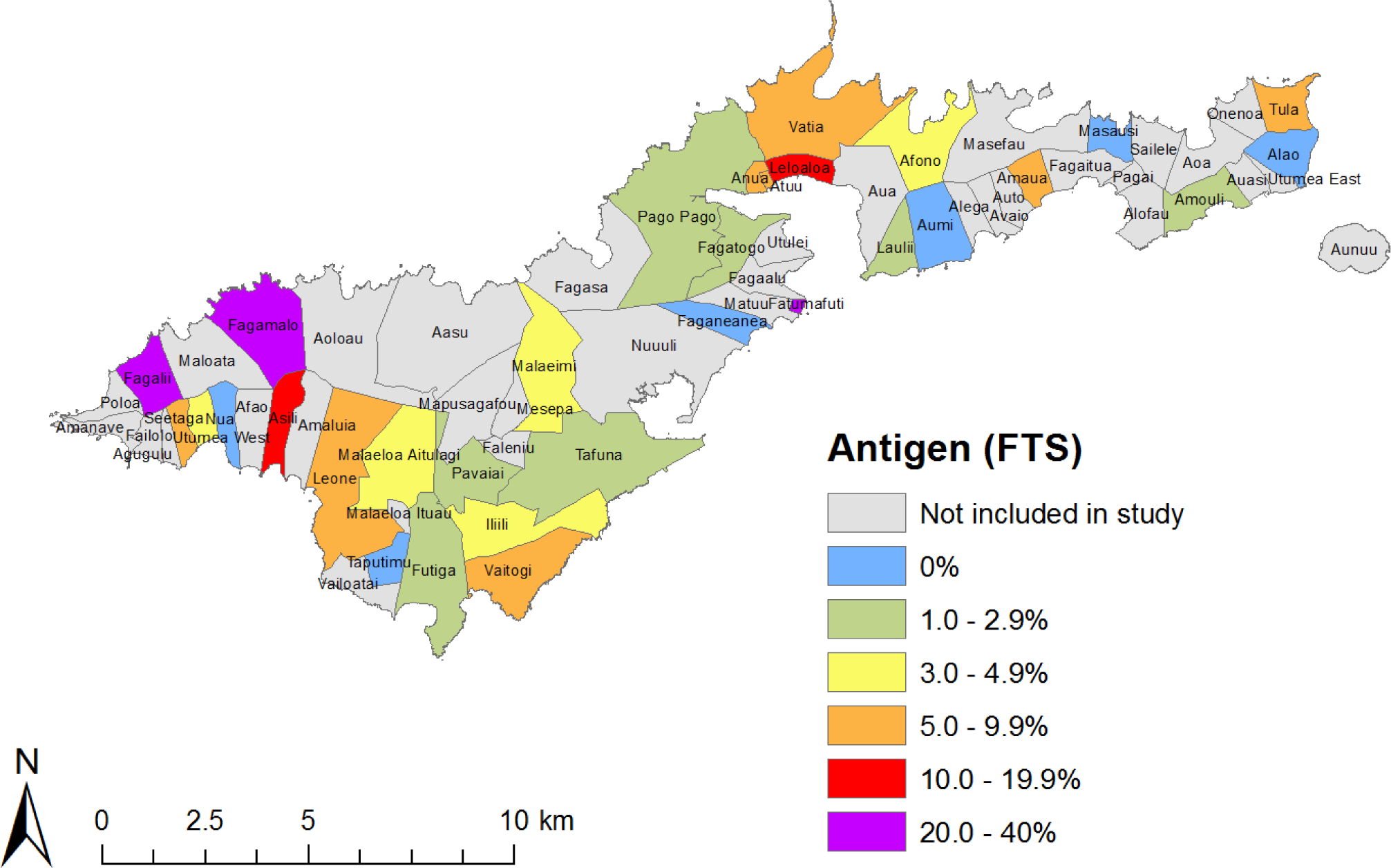
Adjusted seroprevalence by village in community survey (age ≥8 years) of randomly selected villages and previously identified hotspots, American Samoa 2016.

**Figure 6.**
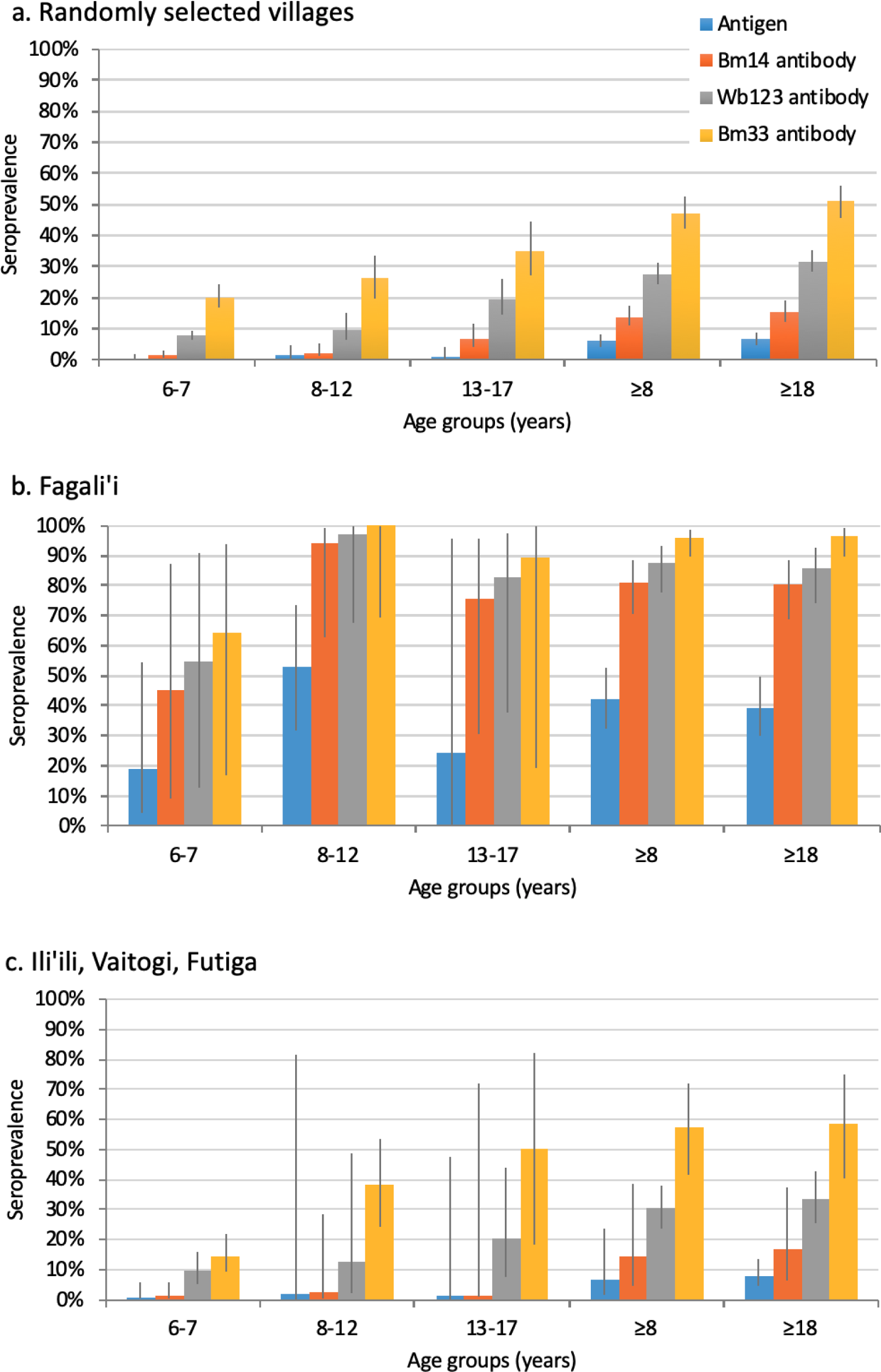
Adjusted seroprevalence (and 95% CI) of Ag and antibodies by age groups for a) randomly selected communities, b) Fagali’i, and c) Ili’ili/Vaitogi/Futiga.

#### A3. Residents of previously identified hotspots

The 2016 community survey included 86 participants aged ≥8 years from Fagali’i (age 8-77 years), and 624 from Ili’ili/Vaitogi/Futiga (age 8-90 years). Adjusted Ag prevalence for Fagali’i and Ili’ili/Vaitogi/Futiga were 42.3% (95% CI 32.7-52.5%) and 6.5% (95% CI 1.5-23.6%), respectively. Of the 27 Mf slides examined from Fagali’i, 12 (44.4%) were Mf-positive, and of the 28 slides examined from Ili’ili/Vaitogi/Futiga, 9 (32.1%) were Mf-positive. Figure 6 shows that the seroprevalence of Ag and antibodies by age groups for residents in the two hotspots, compared to randomly selected villages.

TAS-3 also included 139 children from the two hotspots, including seven from Fagali’i, and 132 from Ili’ili/Vaitogi/Futiga. Of the nine Ag-positive children identified in TAS-3, two were residents of Fagali’i and one was a resident of Ili’ili/Vaitogi/Futiga. Ag prevalence of TAS-3 children in these two hotspots were 18.8% (95% CI 4.3-54.7%) and 0.9% (95% CI 0.1-6.0%), respectively.

#### A4. Household members of Ag-positive children from TAS-3

The nine index children identified in TAS-3 lived in nine households in six different villages. A total of 58 household members were surveyed from the nine households. Of these, 32 (55.2%) were female, and 52 (89.7%) were aged ≥8 years, representing 80% of the reported 65 household members aged ≥8 years who lived in these households. Of the 58 household members tested, 12 (20.7%, 95% CI 11.2-33.4%) were Ag-positive; Ag prevalence was 0% in children aged <8 years (n=6), 27.8% (95% CI 9.7-53.5%) in those aged 8-17 years (n=18), and 20.6% (95% CI 8.7-37.9%) in those aged ≥18 years (n=34). Microfilaria slides were available for 9 of the 12 Ag-positive household members, and two of these (22.2%) were Mf-positive.

At least one Ag-positive individual (in addition to the index child) was identified from five (55.6%) of the nine index households, and more than one additional Ag-positive person (range 2-5) was identified in three (33.3%) of these households. In comparison, the 2016 community survey of randomly selected villages identified Ag-positive persons from 79 (11.1%) of the 711 households, and more than one Ag-positive person (range 2-4) in only 18 (2.5%) of the households (Figure 7).

**Figure 7.**
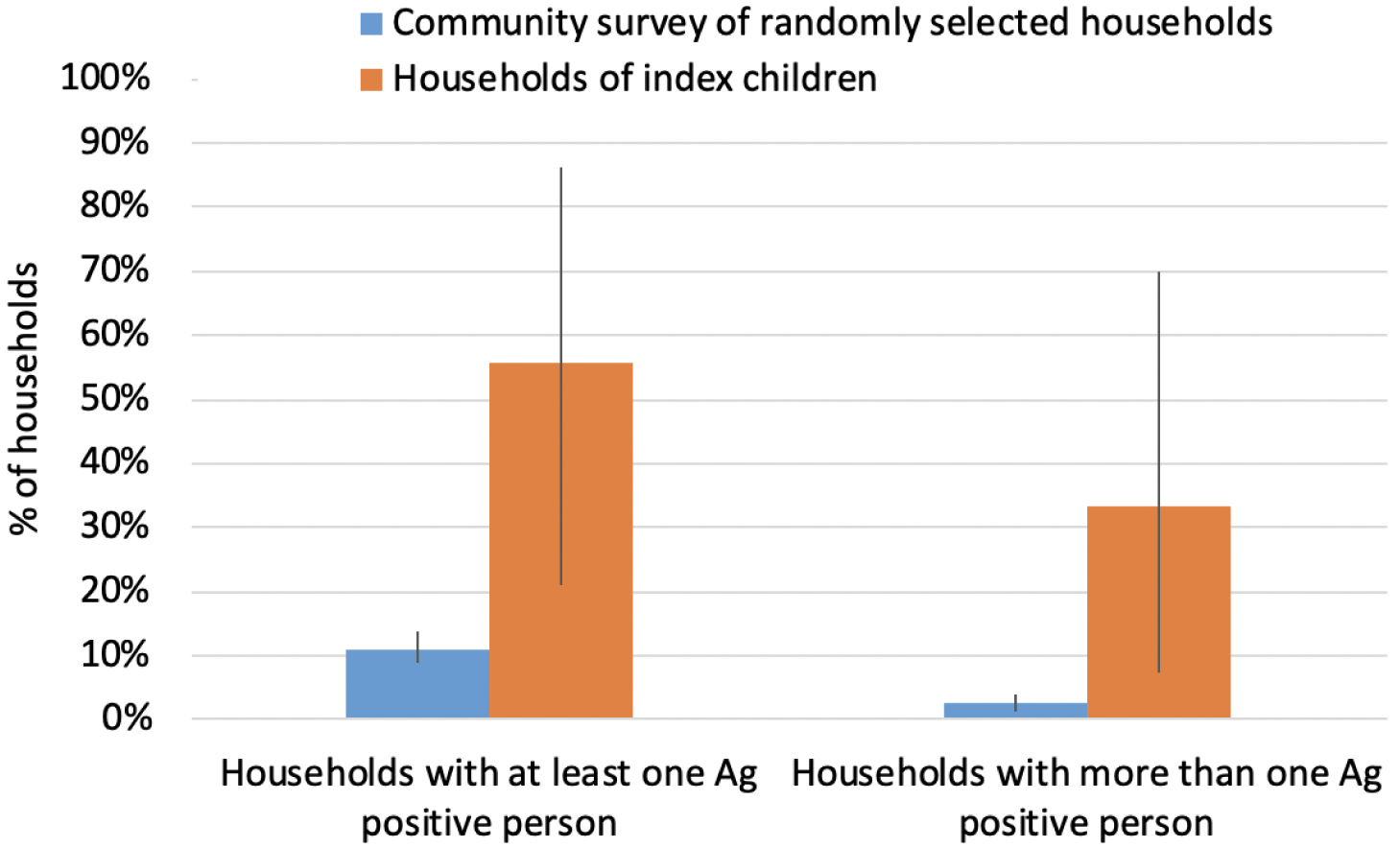
Percentage of households of index children versus randomly selected households in community survey where i) at least one antigen-positive person was identified, and ii) more than one antigen-positive person was identified.

Of the six index villages, four (Vaitogi, Pago Pago, Fagatogo, and Tafuna) were part of the randomly selected villages for the community survey in 2016. One of these (Vaitogi) was also part of a previously identified transmission hotspot (3, 10) and the fifth village (Fagali’i) was purposefully included because previous studies identified it as a transmission hotspot (3, 4). The sixth village (Faga’alu) was not surveyed in 2016, so no village-level seroprevalence data were available. Figure 8 shows that seroprevalence of Ag and antibodies were significantly higher in index households (based on 5 of the 6 villages where community-level data were available) compared to randomly selected households in index villages or in randomly selected villages.

**Figure 8.**
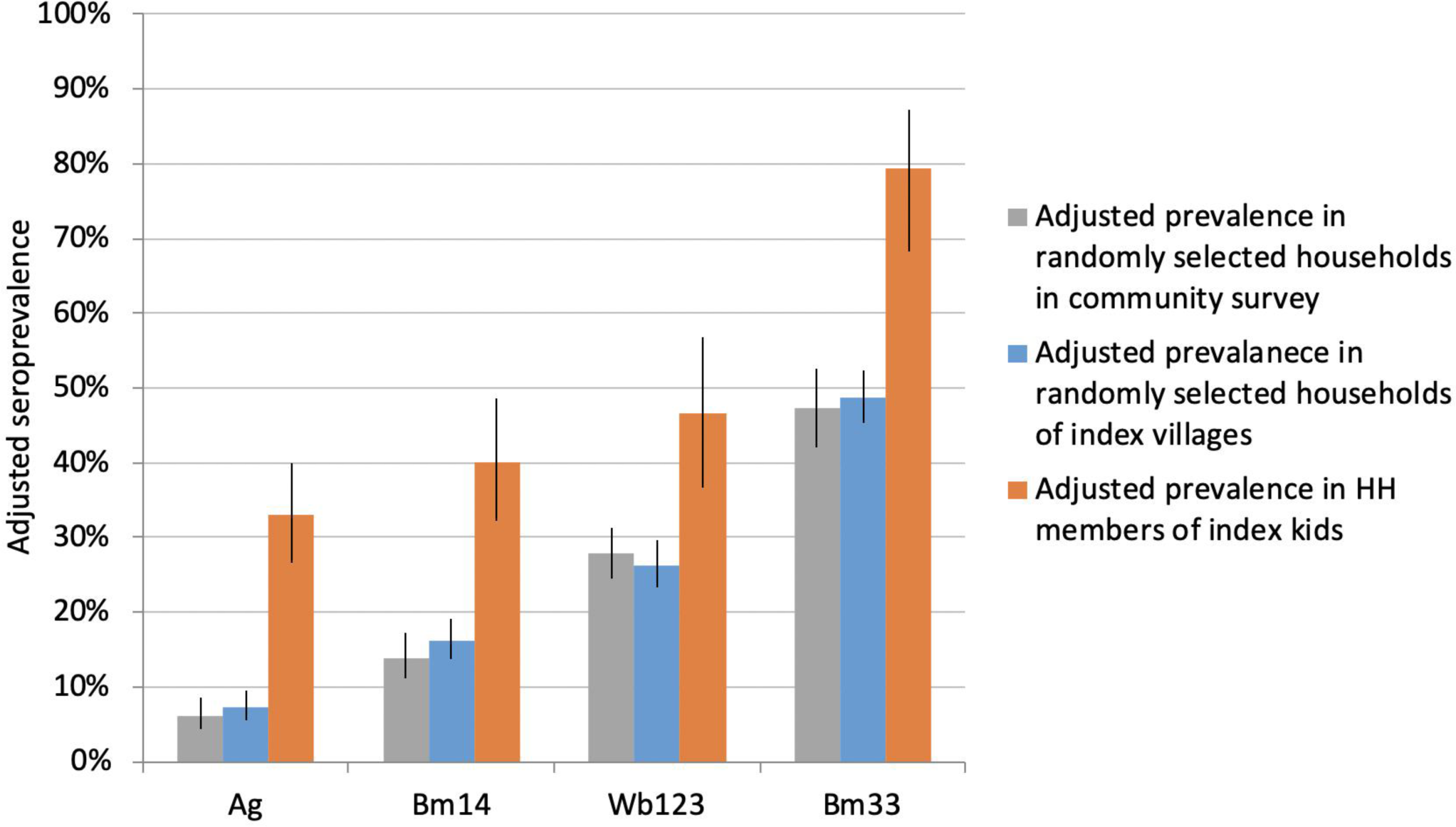
Adjusted seroprevalence of Ag and antibodies in i) randomly selected households in community survey, ii) randomly selected households in index villages, and iii) household members of index children. To enable direct comparisons, results for all groups were limited to participants aged ≥8 years.

### B. Risk factors analysis and logistic regression models

The prevalence of demographic and behavioural factors, and their association with positive seromarkers and Mf are summarised in Tables 1 (for Ag and Mf) and 2 (for antibodies). For all seromarkers and Mf, males, older age groups, and residents of Fagali’i were more likely to be positive, while indoor workers were less likely to be positive. Those who had travelled to an LF-endemic area in the past 12 months (96.2% to Samoa) were more likely to be seropositive for Bm33 antibody, but not the other seromarkers or Mf. The minority (∼16%) who had travelled to non-LF endemic areas in the past 12 months (most commonly to mainland USA, Hawaii, New Zealand, and Australia) were less likely to be seropositive for Ag and antibodies; this result may be confounded by socioeconomic status rather than any causal link. The number of years lived in American Samoa was not associated with Ag or antibody seropositivity, but those who had lived in American Samoa for over 10 years were more likely to be Mf-positive. Participants who reported taking MDA in the past were more likely to be seropositive for Ag and antibodies, but this was not significant once the results were controlled for age in the multivariable analyses.

**Table 1.**
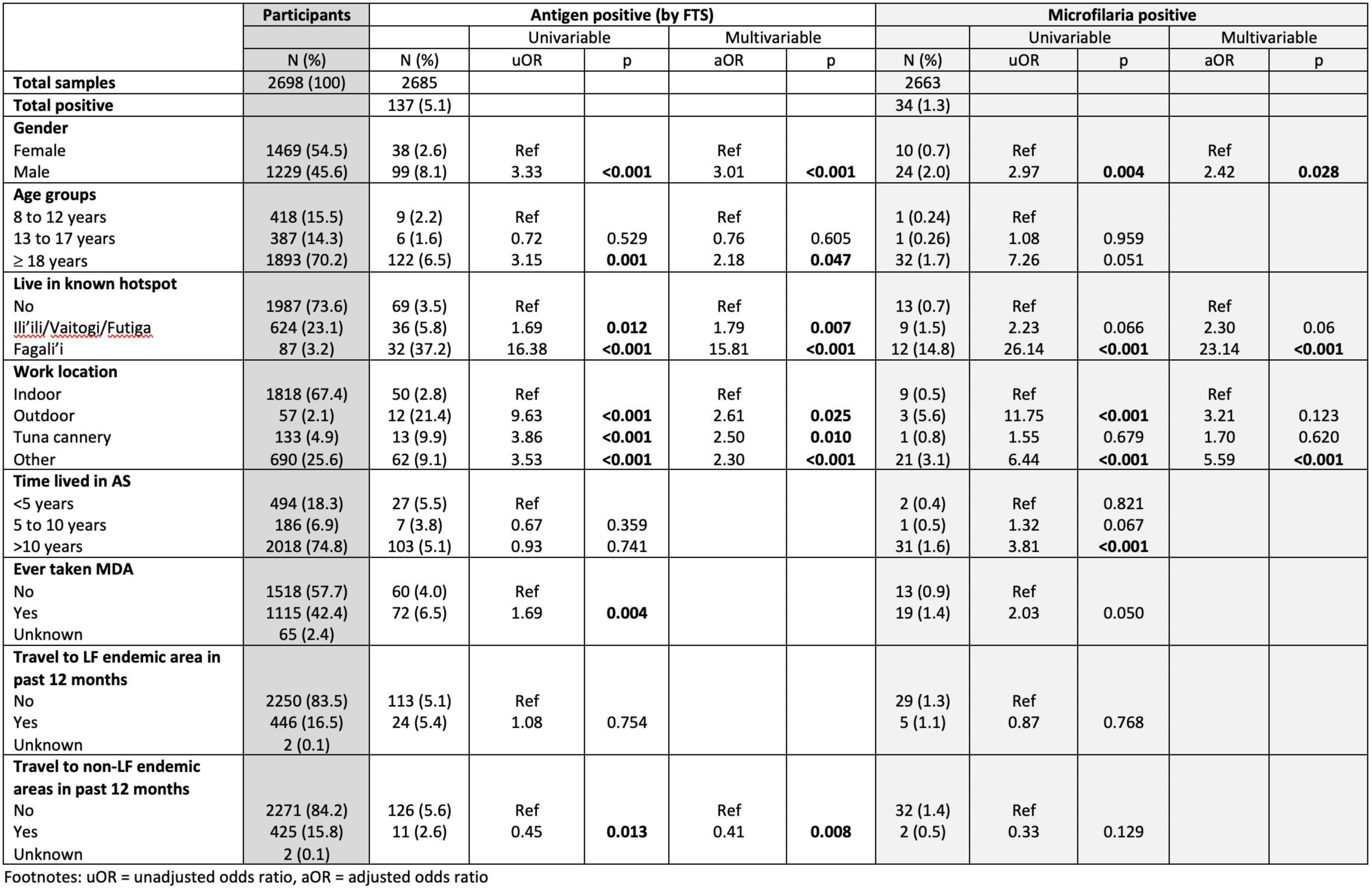
Summary of demographic and behavioural factors, and their associations with antigen-positive and microfilaria-positive results on univariable and multivariable logistic regression.

**Table 2.**
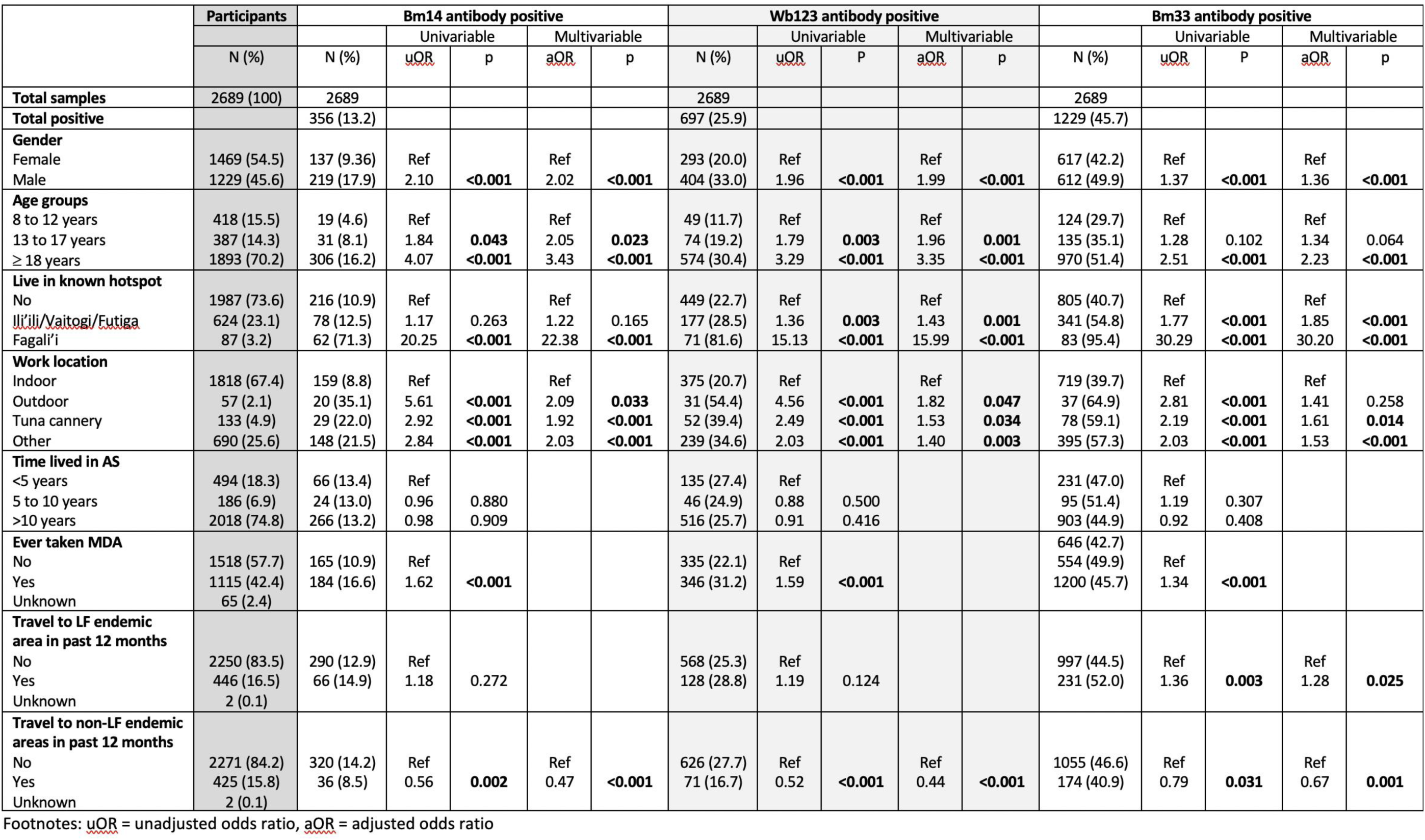
Summary of demographic and behavioural factors, and their associations with positive Bm14, Wb123, and Bm33 antibodies on univariable and multivariable logistic regression.

### C. Intra-cluster correlation (ICC)

ICCs were higher at the household than PSU levels for Mf and all seromarkers, and highest for Mf, followed by Ag, Bm14 Ab, Wb123 Ab, and Bm33 Ab (Figure 9).

**Figure 9.**
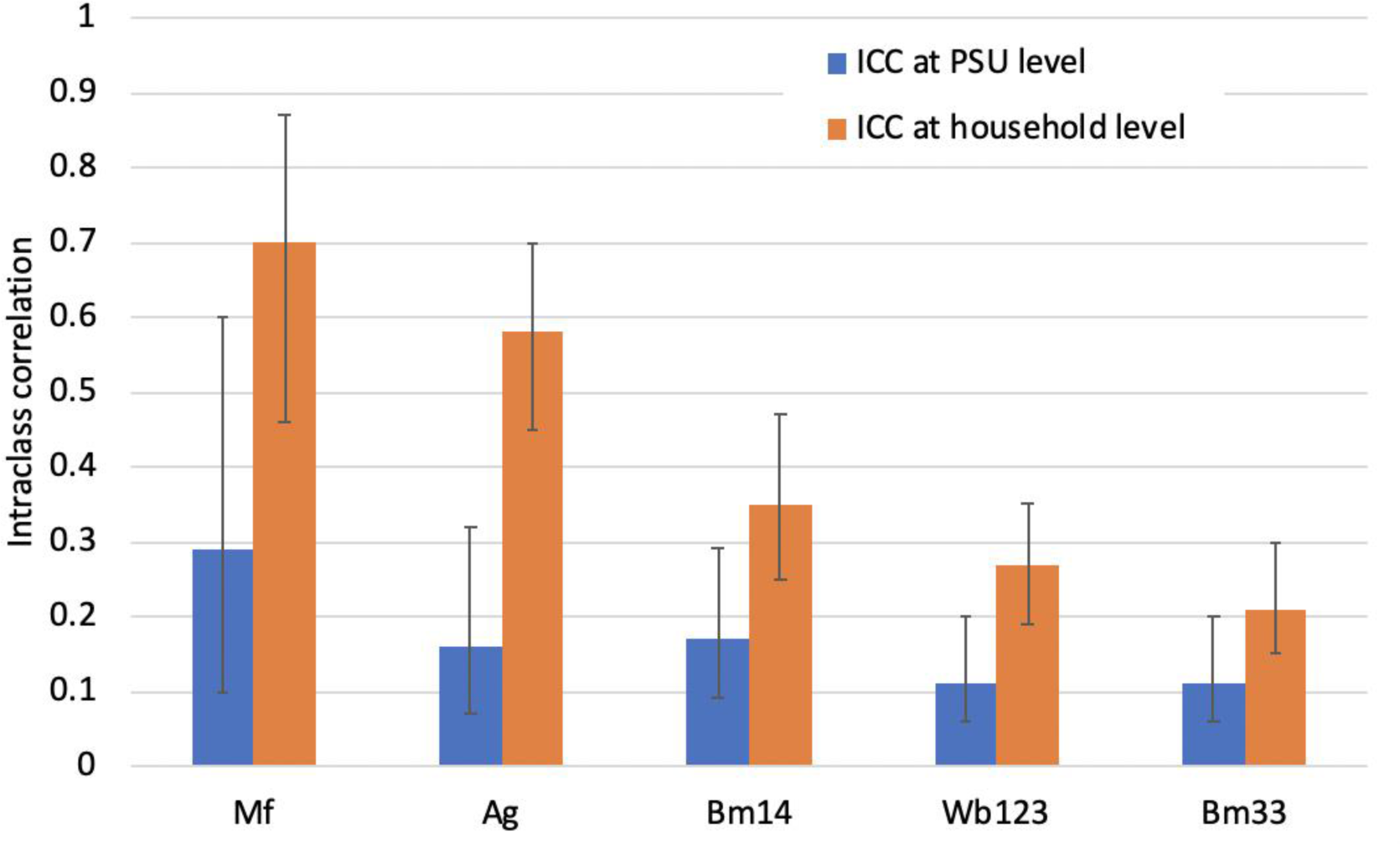
Intra-cluster correlation (ICC) and 95% CI at PSU and household levels for positive results for microfilaria, Ag, and antibodies (Bm14, Wb123, and Bm33).

### D. Number needed to test (NNTest)

NNTest^av^, NNTest^50^, and NNTest^95^ were calculated for subgroups based on survey strategy (population representative surveys, hotspots, household members of Ag-positive children) and age groups, and summarised in Figure 10. For all subgroups, all three NNTest measures were highest for Ag, followed by Bm14, Wb123, and Bm33 antibodies. NNTest also generally decreased as the age of the target population increased. Population representative surveys such as TAS and community surveys were associated with significantly higher NNTest (i.e. less efficient) than more targeted surveillance strategies.

**Figure 10.**
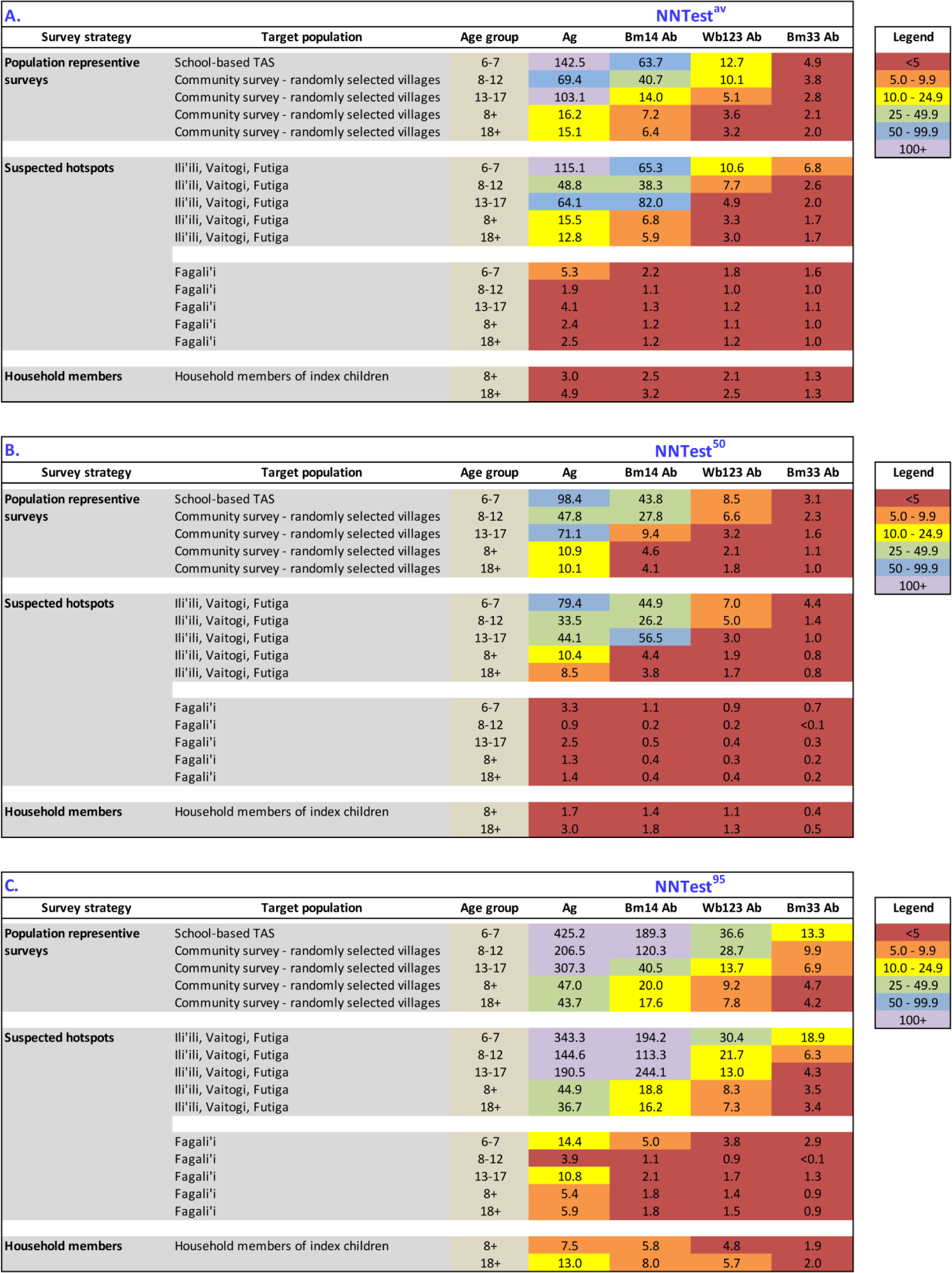
Number need to test for different surveillance strategies, target populations, and age groups using antigen and Bm14, Wb123, and Bm33 antibodies. A) Average number needed to test to identify one positive person (NNTest^av^), B) Number needed to test to provide a 50% chance of identifying at least one seropositive person (NNTest^50^), and C) Number needed to test to provide a 95% chance of identifying at least one seropositive person (NNTest^95^).

If targeted surveillance was conducted in households of index children, the NNTest^av^ was ≤3 regardless of the diagnostic test used. In these households, where additional Ag-positive individuals were identified from five of nine households, an average of only 1.8 households needed to be surveyed in order to find at least one Ag-positive person, compared to an average of 9.0 households (5 times as many) for randomly selected households.

Figure 11 shows the probability of detecting at least one positive case based on selected survey strategies with different levels of NNTest^av^ and sample size (assuming that positive cases are independent events). S4 Figure shows the probability of detecting at least one positive case based on sample size and NNTest^av^ of 5, 10, 20, 50, 100, and 150 (assuming that positive cases are independent events).

**Figure 11.**
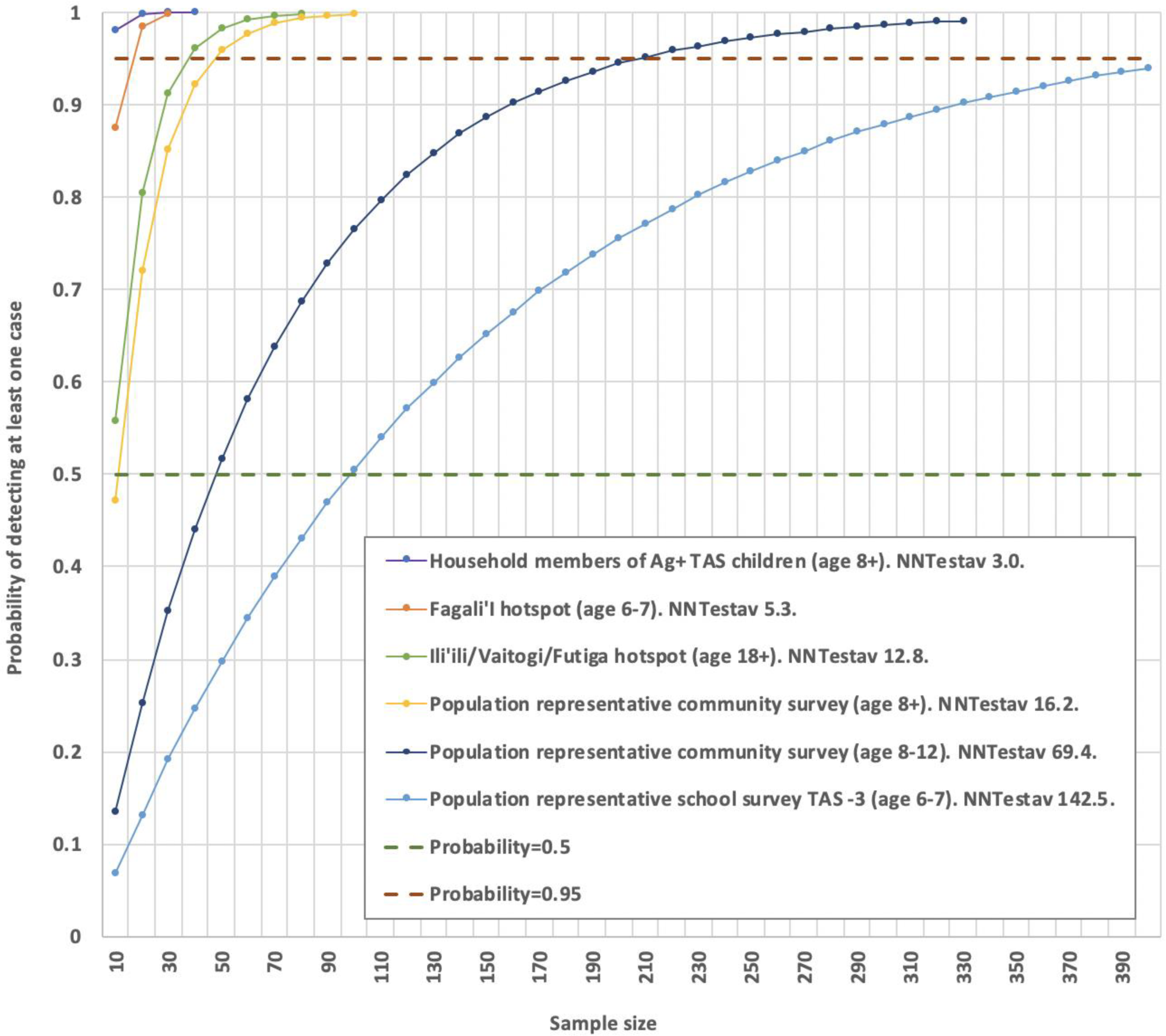
Probability of detecting at least one positive case based on sample size and selected survey strategies with different levels of NNTest^av^ (assumes that positive cases are independent events).

## DISCUSSION

For LF elimination programmes, the goals of surveillance are to identify any remaining Ag-positive people and residual transmission hotspots, and to ideally pick up signals of resurgence at an early stage. Our study demonstrated a number of potential strategies for strengthening surveillance and improving the likelihood of achieving these goals. In the post-MDA setting, the ability of surveillance strategies to efficiently identify any signals of residual transmission and/or resurgence is critical but challenging, particularly if infected persons are not randomly distributed within a population. Our study quantified the degree to which post-MDA surveillance strategies that specifically target high-risk populations and locations can improve sampling efficiency at identifying such signals, and provided evidence to suggest that these strategies should be considered together with population representative surveys such as TAS. Compared to current recommendations for TAS in 6-7 year-old children, we found that the NNTest was significantly lower if testing older children and/or adults, conducting targeted surveillance (of previously identified hotspots and households of index children), using antibodies, or a combination of these strategies. Although these strategies are more efficient than TAS for identifying infected persons, they alone will not provide accurate estimates of population-level seroprevalence. Therefore, population representative surveys such as TAS will be required to estimate true prevalence as well as provide a starting point to inform the targeted strategies.

Higher seroprevalence of Ag and antibodies in older age groups is a common finding in LF surveys across the world. The current TAS recommendation for testing 6-7 year-old children is based on the rationale that detection of infections in young children (who were born after MDA had started) provides evidence of ongoing transmission. Also, conducting school-based surveys of children is generally logistically simpler and cheaper than community-based household surveys (5). However, the low prevalence of infection in young children means that the NNTest is significantly higher compared to testing older age groups, resulting in reduced efficiency and cost-effectiveness of surveys, as well as a higher chance of missing early signals of resurgence. While positive Ag in children represent more recent infections, active infections in adults are also important indicators of remaining reservoirs of infection that should be addressed. Our study showed that population representative surveys of older children have the potential to reduce NNTest, and cost-effectiveness could be maintained by conducting the surveys at schools in areas where school attendance rates are high, e.g. testing age 8-12 year-olds in elementary schools, or age 13-17 year-olds at secondary school. Furthermore, testing 8-12 year-olds in TAS-2, TAS-3, and beyond is consistent with the rationale behind TAS, i.e targeting age groups who have lived their entire lives during and/or after MDA, so any positive cases would indicate incident infection and therefore ongoing transmission. School-based TAS of older children should therefore be considered, and together with adjunct surveillance strategies discussed above, could potentially optimise the detection of residual infections while maintaining cost-effectiveness. Population representative surveys in communities have the potential to further reduce NNTest (because adults can be included) and provide more detailed epidemiological information than school-based surveys, but are logistically more complex and more expensive (16).

Our findings provide strong evidence to support targeted surveillance and/or empirical treatment of household members of infected persons. Household members of index children had significantly higher risk of infection than any other groups, with an NNTest^av^ of five or less regardless of the serological marker used. Higher infection prevalence in household members compared to community members of index children also indicates that infections are intensely clustered at the household level. All infected persons should therefore be considered as potential sentinels of microfoci of infection, regardless of how they were identified. Based on current TAS guidelines, household members of index children identified through TAS should be tested and/or treated, but this is not always done. Considering that NNtest^av^ was <5 for household members of Ag-positive persons, and more than half the index households had at least one Ag-positive person, testing/treating household members would be a very cost effective public health strategy for reducing reservoirs of infection, and also clinically important for reducing the risk of long-term morbidity in individuals. If future surveillance recommendations include other strategies (e.g. testing older children at school, opportunistic testing of adult workers (4, 22)), testing/treating all household members of seropositive persons should also be considered. Population representative survey strategies that reduce NNTest can further improve the efficiency of identifying sentinels that could lead us to more households with high prevalence of infection.

Targeted surveillance of known hotspots is another potential strategy for reducing NNTest, particularly if older age groups were tested and/or if antibodies were used. In American Samoa, hotspots were identified by previous studies conducted in 2010 and 2014. In 2016, some of these areas again had higher seroprevalence of Ag and antibodies, suggesting ongoing transmission. Currently, there is limited information on the size of transmission hotspots, and whether this differs between places, environmental conditions, vector species, time since MDA, or other factors. Our previous study in American Samoa suggested cluster size of 1.2 to 1.5km, while a study in Haiti found a significantly higher likelihood of being Ag-positive if living within 20 metres of index cases (11). Further studies are required to determine the factors that influence cluster size, the risk of infection in near neighbours of index cases, whether targeted surveillance of these neighbours should be recommended, and how large the surveillance buffer should be.

Based on our finding of intense household-level clustering, surveillance of near neighbours of index households could also be considered. Considering the focal nature of residual hotspots, geographically driven surveillance strategies are likely to improve precision and reduce cost. However, improved knowledge about the temporal and spatial characteristics of hotspots will be required to optimise these strategies. Geostatistical and other spatially-explicit models may be able to predict hotspots based on known locations of infections, environmental factors, and social connectivity (30-33). In the future, these models could potentially further reduce NNTest by directing the surveillance team to locations where infected persons are most likely to be found.

Our study found that outdoor workers were more likely to be seropositive for Ag and antibodies. We did not include work location in our NNTest matrices, but it is possible to include this and other risk factors, especially if it is feasible to develop targeted surveillance strategies for these subpopulations. In this study, the number of years lived in American Samoa was not associated with seroprevalence of Ag or antibodies. This is in contrast to our previous study using blood samples collected in 2010 (two years after the last MDA), but consistent with our study in 2014 (six years after the last MDA). One possible explanation for the differences in findings between studies is that in a setting of resurgence, some ten years after MDA, previous residential history (and therefore timing and likelihood of MDA participation) gradually makes less difference as time goes by.

Currently, Ag is the key seromarker used for LF surveillance, detected using the Alere FTS rapid diagnostic test (RDT). Considering that antibody prevalence is typically higher than Ag prevalence, using antibodies for surveillance (possibly together with Ag) could reduce NNTest. Studies in children in Samoa and Sri Lanka found that antibody was more sensitive than Ag for detecting residual transmission of LF in the post-MDA setting (34, 35). The two main barriers to using antibodies for surveillance are the poor understanding of the changes in antibody patterns over time, and the lack of an RDT that can be used in field surveys. Antigen detected by tests such as FTS (36, 37) is produced by adult filarial worms in the lymphatic system and circulates in the peripheral blood during the adult worm’s lifespan and for an unknown period thereafter. The Wb123 antibody was identified from a library generated from L3 larval stages of *W. bancrofti* (38), and is detectable in the early stages of infection, but also persists for long periods during and after infection in adults (39). The Bm14 antibody was identified from a cDNA library screened using sera from microfilariaemic people (40, 41), and may persist for many years post-infection. Bm33 antibody was also identified in a *B.malayi* cDNA library as a major cross-reacting immunogen in *W. bancrofti* (42). The duration of persistence of Bm14 and Bm33 antibodies is long (many years) but not well known. The discordance between Ag and antibodies results in young children indicate that serological patterns in recently acquired infections are complex, and each serological marker most likely provides different information about infection and immune status. Further studies are needed to understand how serological patterns of LF antigen and antibodies evolve over time, the sensitivity and specificity of antibodies as indicators of recent or active infection, and their utility as surveillance tools.

The NNTest results presented in this paper reflect the epidemiology of LF in American Samoa. For other locations, NNTest might differ depending on infection prevalence, the degree of community and household level clustering (ICCs), and the characteristics of hotspots. For future cluster surveys, sample size calculations should adjust for design effect at both the PSU and household levels. Considering that ICC is higher for Ag than antibodies, the design effect will be also higher. For strategies with already high NNTest, the need to multiply this by a large design effect could significantly further increase the primary and/or secondary sampling units required.

Other strategies that could improve the cost-effectiveness of post-MDA surveillance include integrating surveillance with other diseases or public health programs, and opportunistic screening of large populations such as workers, military recruits and blood donors (4, 43). Our previous work in American Samoa demonstrated that community-based survey of older age groups may provide earlier, more sensitive, and/or more geographically precise signals of ongoing transmission than TAS (5), but was logistically more challenging and more expensive. Screening of adults could therefore reduce NNTest (43), and if conducted through an efficient sampling strategy (e.g. opportunistic screening at workplaces or clinics (4)), it is potentially possible to achieve the desirable combination of low NNTest and low cost. In American Samoa (16) and Sri Lanka (8), molecular xenomonitoring of mosquitoes has also been shown to potentially provide earlier and more sensitive signals of transmission than TAS, but requires high level entomological expertise in both mosquito identification and molecular testing which may be beyond the reach of many countries.

Taking together findings from our current and previous studies, an approach for improving effectiveness and efficiency of surveillance could be a multi-stage test-and-treat strategy for identifying and eliminating residual foci of transmission. For example, a multi-stage operational framework for post-MDA surveillance could start with population representative sampling that takes into account the NNTest for different target age groups, and operational costs and logistics for different survey sites (e.g. community, school, work sites). Opportunistic screening, e.g. at work sites, could also be considered. The next step would be a more intensive targeted sampling of any high-risk groups (including household members of infected persons) and high prevalence locations and hotspots (Figure 12). Targeted sampling should consider all relevant data from the population representative survey and make the most use of any clues about the demographics and locations of high-risk persons. For example, starting with a school-based survey of older children, followed by active targeted surveillance or ‘contact tracing’ of index households, and more intensive surveillance of hotspots. For both the population representative and targeted sampling stages, programs need to decide on the level of uncertainty that they are prepared to accept, and balance this against the sample size (and cost) required. For example, in populations with low NNTest (e.g. in hotspots), a small increase in sample size is associated with a steep increase in the probability of identifying at least one case; targeted sampling in these areas would therefore be highly likely to identify other infected persons and provide further sentinels. In contrast, in populations with very high NNTest (e.g. TAS of 6-7 year-olds), a relatively large increase in sample size is required to increase the probability of detecting one case, particularly as the probability approaches one (Figures 11 and S4).

**Figure 12.**
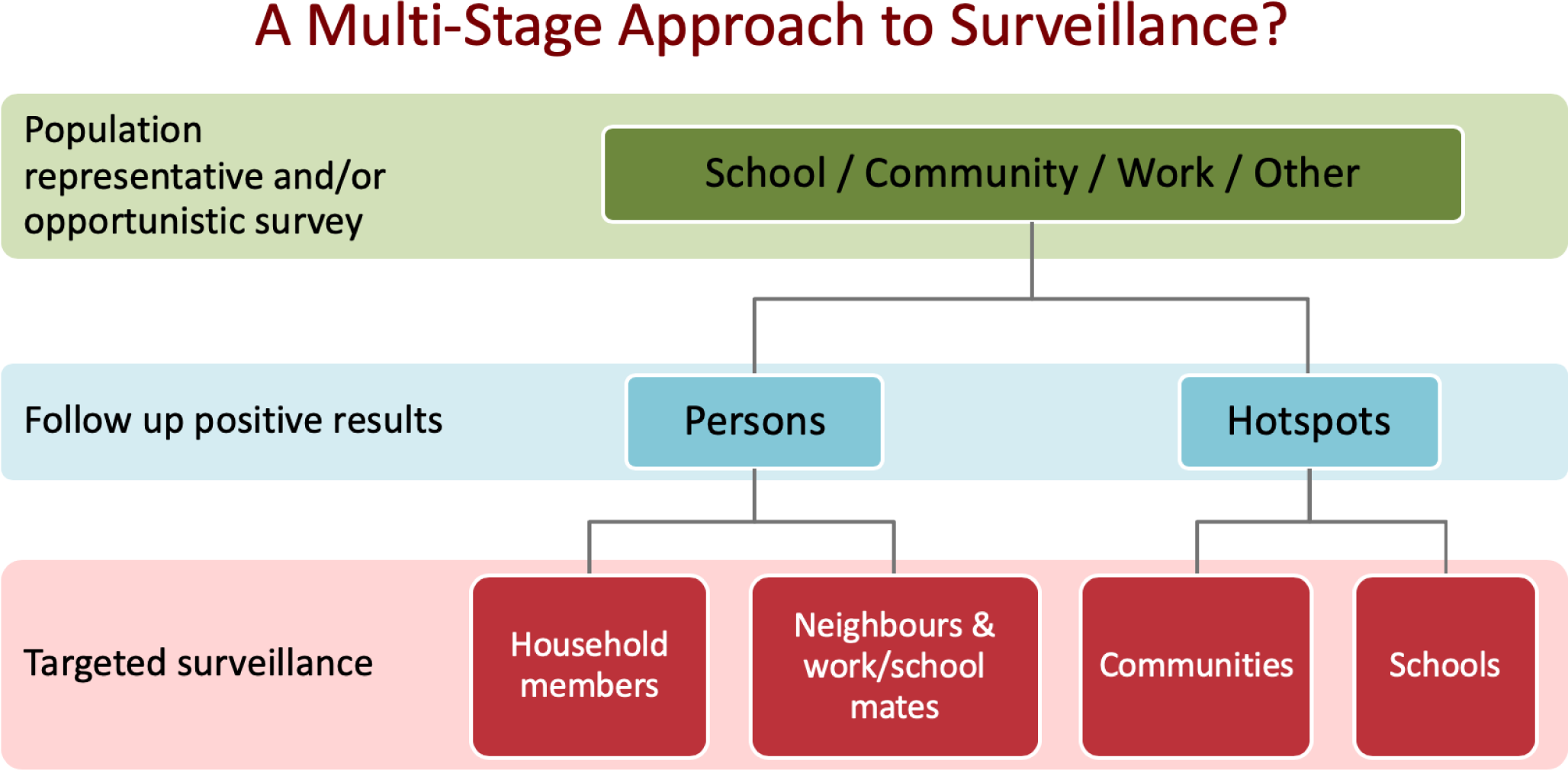
Potential framework for a multi-stage surveillance strategy for LF elimination programs

The strengths of our study include high-resolution data that enabled linkage of individuals by households and communities, testing and surveying household members of index children, and knowledge of previously identified hotspots which we could further assess. By conducting a school-based TAS and a community survey in parallel, we were also able to accurately compare results between the two survey methods. The results should be considered in light of the study’s limitations. The study was designed to detect differences between 6-7 year-olds and those aged 8 years and older, but not the smaller age bands of 8-12 years, 13-17 years, and ≥18 years. Despite this, the significant difference in seroprevalence between the different age groups allowed detailed comparisons. Similarly, the study was not powered to detect differences between PSUs, but significantly higher Ag and antibody prevalence were noted in the previous identified hotspots. The study was conducted in American Samoa, a remote tropical island location where LF is caused by *Wuchereria bancrofti* and transmitted by *Aedes* mosquitoes; the concepts explored in our study could be applied to other locations, but the quantitative results might differ in other settings. Another limitation of surveillance strategies that rely on population-representative surveys (e.g. cluster sampling) is that they may miss residual infections and hotspots altogether. Therefore, the overall success of any targeted surveillance program and/or test-and-treat program depends on the sensitivity of the initial survey(s). Although TAS in American Samoa included all elementary schools, this is not the case for most countries, where only a small proportion of schools or communities are tested. Improving the representativeness and sensitivity of the initial survey is therefore crucial for overall success, e.g. geospatial models to guide adaptive sampling of clusters, and more strategic sampling in TAS-2 and TAS-3 to ensure greater representativeness of the entire evaluation unit. Currently, there is limited evidence on the Ag or antibody thresholds for conducting targeted MDA at a broader population level; this information is important for ensuring that undetected infections are also treated.

## CONCLUSION

Historical evidence demonstrates that LF can resurge in the Pacific islands even when infection rates have dropped to very low levels (44, 45). Possible reasons include the combination of efficient vectors, highly mobile population (31), outdoor lifestyle, MDA coverage/participation rates (21), and remote/isolated populations that make drug distribution and surveillance a challenge. Public health resources are usually limited and need to be shared amongst many health priorities. Taking these factors together, long-term success of the LF elimination programme in the Pacific Islands (and most probably elsewhere) will require highly effective and efficient surveillance strategies that maximise the likelihood of identifying residual foci of transmission and detecting early signals of any resurgence.

## Data Availability

The datasets generated and analysed during the present study are available from the corresponding author upon reasonable request.

## Acknowledgements

We would like to acknowledge the hard work of all our field team members, particularly Ms Paeae Tufono, Ms Ledonna Pule, Ms Fitilagi Tagiola, Ms Susana Lin, Ms Meliame Tufono, Ms Nalini Lata, Mr Jason Tufele and Ms Catherine Montablo. We would also like to thank Ms Mary Matai’a of the American Samoa Department of Health for her assistance with logistics and laboratory testing of specimens. We also thank Dr Mark Schmaedick at the American Samoa Community College for generously allowing us to use his laboratories. We are also grateful to all the school principals and teachers, and village mayors and chiefs for their assistance in conducting the fieldwork. The findings and conclusions in this paper are those of the authors and do not necessarily represent the official position of the CDC.

## Supporting Information Captions

S1 Appendix. Methods used to calculate seroprevalence adjusted for age, sex, and survey design. S2 Table. Probability of selection and post-stratification weights of different subgroups.

S3 Table. Adjustments used for different subgroups.

S4 Figure. Probability of detecting at least one positive case based on sample size and NNTestav of 5, 10, 20, 50, 100, and 150 (assumes that positive cases are independent events).

S5 Checklist. STROBE checklist for cross-sectional studies.

## References

1. Global Programme to Eliminate Lymphatic Filariasis: Progress Report, 2017. Wkly Epidemiol Rec. 2018;93(44):589–604.

2. World Health Organization. Monitoring and Epidemiological Assessment of Mass Drug Administration in the Global Programme to Eliminate Lymphatic Filariasis: A Manual for National Elimination Programmes. 2011. [Cited 2020 Feb 28]. Available from: http://whqlibdoc.who.int/publications/2011/9789241501484_eng.pdf.

3. Lau CL, Won, KY, Becker, L, Soares Magalhaes, RJ, Fuimaono, S, Melrose, W, Lammie, PJ, Graves, PM. Seroprevalence and Spatial Epidemiology of Lymphatic Filariasis in American Samoa after Successful Mass Drug Administration. PLoS Negl Trop Dis. 2014;8(11):e3297.

4. Lau CL, Sheridan, S, Ryan, S, Roineau, M, Andreosso, A, Fuimaono, S, Tufa, J, Graves, PM. Detecting and Confirming Residual Hotspots of Lymphatic Filariasis Transmission in American Samoa 8 Years after Stopping Mass Drug Administration. PLoS Negl Trop Dis. 2017;11(9):e0005914.

5. Sheel M, Sheridan, S, Gass, K, Won, K, Fuimaono, S, Kirk, M, Gonzales, A, Hedtke, SM, Graves, PM, Lau, CL. Identifying Residual Transmission of Lymphatic Filariasis after Mass Drug Administration: Comparing School-Based Versus Community-Based Surveillance - American Samoa, 2016. PLoS Negl Trop Dis. 2018;12(7):e0006583.

6. Coutts SP, King, JD, Pa’au, M, Fuimaono, S, Roth, J, King, MR, Lammie, PJ, Lau, CL, Graves, PM. Prevalence and Risk Factors Associated with Lymphatic Filariasis in American Samoa after Mass Drug Administration. Trop Med Health. 2017;45:22.

7. Joseph H, Moloney, J, Maiava, F, McClintock, S, Lammie, P, Melrose, W. First Evidence of Spatial Clustering of Lymphatic Filariasis in an Aedes Polynesiensis Endemic Area. Acta Trop. 2011;120 Suppl 1:S39–47.

8. Rao RU, Samarasekera, SD, Nagodavithana, KC, Goss, CW, Punchihewa, MW, Dassanayaka, TDM, Ranasinghe, USB, Mendis, D, Weil, GJ. Comprehensive Assessment of a Hotspot with Persistent Bancroftian Filariasis in Coastal Sri Lanka. Am J Trop Med Hyg. 2018.

9. Srividya A, Subramanian, S, Sadanandane, C, Vasuki, V, Jambulingam, P. Determinants of Transmission Hotspots and Filarial Infection in Households after Eight Rounds of Mass Drug Administration in India. Trop Med Int Health. 2018;23(11):1251–8.

10. Washington CH, Radday, J, Streit, TG, Boyd, HA, Beach, MJ, Addiss, DG, Lovince, R, Lovegrove, MC, Lafontant, JG, Lammie, PJ, Hightower, AW. Spatial Clustering of Filarial Transmission before and after a Mass Drug Administration in a Setting of Low Infection Prevalence. Filaria J. 2004;3(1):3.

11. Drexler N, Washington, CH, Lovegrove, M, Grady, C, Milord, MD, Streit, T, Lammie, P. Secondary Mapping of Lymphatic Filariasis in Haiti-Definition of Transmission Foci in Low-Prevalence Settings. PLoS Negl Trop Dis. 2012;6(10):e1807.

12. Lammie PJ, Eberhard, ML, Addiss, DG, Won, KY, Beau de Rochars, M, Direny, AN, Milord, MD, Lafontant, JG, Streit, TG. Translating Research into Reality: Elimination of Lymphatic Filariasis from Haiti. Am J Trop Med Hyg. 2017;97(4_Suppl):71-5.

13. World Health Organization. Lymphatic Filariasis - Research 2016 [Cited 2020 Feb 28]. Available from: http://www.who.int/lymphatic_filariasis/research/en/.

14. Hamlin KL, Moss, DM, Priest, JW, Roberts, J, Kubofcik, J, Gass, K, Streit, TG, Nutman, TB, Eberhard, ML, Lammie, PJ. Longitudinal Monitoring of the Development of Antifilarial Antibodies and Acquisition of Wuchereria Bancrofti in a Highly Endemic Area of Haiti. PLoS Negl Trop Dis. 2012;6(12):e1941.

15. Won KY, Robinson, K, Hamlin, KL, Tufa, J, Seespesara, M, Wiegand, RE, Gass, K, Kubofcik, J, Nutman, TB, Lammie, PJ, Fuimaono, S. Comparison of Antigen and Antibody Responses in Repeat Lymphatic Filariasis Transmission Assessment Surveys in American Samoa. PLoS Negl Trop Dis. 2018;12(3):e0006347.

16. Lau CL, Won, KY, Lammie, PJ, Graves, PM. Lymphatic Filariasis Elimination in American Samoa: Evaluation of Molecular Xenomonitoring as a Surveillance Tool in the Endgame. PLoS Negl Trop Dis. 2016;10(11):e0005108.

17. Pilotte N, Unnasch, TR, Williams, SA. The Current Status of Molecular Xenomonitoring for Lymphatic Filariasis and Onchocerciasis. Trends Parasitol. 2017;33(10):788–98.

18. Rao RU, Samarasekera, SD, Nagodavithana, KC, Punchihewa, MW, Dassanayaka, TD P KDG, Ford, E, Ranasinghe, US, Henderson, RH, Weil, GJ. Programmatic Use of Molecular Xenomonitoring at the Level of Evaluation Units to Assess Persistence of Lymphatic Filariasis in Sri Lanka. PLoS Negl Trop Dis. 2016;10(5):e0004722.

19. Subramanian S, Jambulingam, P, Chu, BK, Sadanandane, C, Vasuki, V, Srividya, A, Mohideen AbdulKader, MS, Krishnamoorthy, K, Raju, HK, Laney, SJ, Williams, SA, Henderson, RH. Application of a Household-Based Molecular Xenomonitoring Strategy to Evaluate the Lymphatic Filariasis Elimination Program in Tamil Nadu, India. PLoS Negl Trop Dis. 2017;11(4):e0005519.

20. World Health Organization ROftWP, Filariasis., PPtEL. The Pacelf Way: Towards the Elimination of Lymphatic Filariasis from the Pacific, 1999-2005.: Manila: WHO Regional Office for the Western Pacific; 2006. [Cited 2020 Feb 28]. Available from: http://iris.wpro.who.int/handle/10665.1/10966.

21. King JD, Zielinski-Gutierrez, E, Pa’au, M, Lammie, P. Improving Community Participation to Eliminate Lymphatic Filariasis in American Samoa. Acta Trop. 2011;120 Suppl 1:S48–54.

22. Graves PM, Sheridan, S, Fuimaono, S, Lau, CL. Demographic, Socioeconomic and Disease Knowledge Factors, but Not Population Mobility, Associated with Lymphatic Filariasis Infection in Adult Workers in American Samoa in 2014. Parasites and Vectors. 2020; In press.

23. Schmaedick MA, Koppel, AL, Pilotte, N, Torres, M, Williams, SA, Dobson, SL, Lammie, PJ, Won, KY. Molecular Xenomonitoring Using Mosquitoes to Map Lymphatic Filariasis after Mass Drug Administration in American Samoa. PLoS Negl Trop Dis. 2014;8(8):e3087.

24. Rembold CM. Number Needed to Screen: Development of a Statistic for Disease Screening. BMJ. 1998;317(7154):307–12.

25. Cook RJ, Sackett, DL. The Number Needed to Treat: A Clinically Useful Measure of Treatment Effect. BMJ. 1995;310(6977):452–4.

26. American Samoa Department of Commerce. 2014 Statistical Yearbook 2014. [Cited 2020 Feb 28]. Available from: http://doc.as.gov/wp-content/uploads/2016/06/2014-Statistical-Yearbook.pdf.

27. Pavluck A, Chu, B, Mann Flueckiger, R, Ottesen, E. Electronic Data Capture Tools for Global Health Programs: Evolution of Links, an Android-, Web-Based System. PLoS Negl Trop Dis. 2014;8(4):e2654.

28. Marine Cadastre Admin. American Samoa Coastal and Marine Spatial Planning Data Portal 2020. [Cited 2020 Mar 10]. Available from: https://www.arcgis.com/home/item.html?id=7db19f0ac94e4f97abc10711e7f540bc.

29. Thulin M. The Cost of Using Exact Confidence Intervals for a Binomial Proportion. Electronic Journal of Statistics. 2013;8(1).

30. Xu Z, Graves, PM, Lau, CL, Clements, A, Geard, N, Glass, K. Geofil: A Spatially-Explicit Agent-Based Modelling Framework for Predicting the Long-Term Transmission Dynamics of Lymphatic Filariasis in American Samoa. Epidemics. 2019;27:19–27.

31. Xu Z, Lau, CL, Zhou, X, Fuimaono, S, Soares Magalhaes, RJ, Graves, PM. The Extensive Networks of Frequent Population Mobility in the Samoan Islands and Their Implications for Infectious Disease Transmission. Sci Rep. 2018;8(1):10136.

32. Stensgaard AS, Vounatsou, P, Onapa, AW, Utzinger, J, Pedersen, EM, Kristensen, TK, Simonsen, PE. Ecological Drivers of Mansonella Perstans Infection in Uganda and Patterns of Co-Endemicity with Lymphatic Filariasis and Malaria. PLoS Negl Trop Dis. 2016;10(1):e0004319.

33. Slater H, Michael, E. Mapping, Bayesian Geostatistical Analysis and Spatial Prediction of Lymphatic Filariasis Prevalence in Africa. PLoS One. 2013;8(8):e71574.

34. Rao RU, Samarasekera, SD, Nagodavithana, KC, Dassanayaka, TDM, Punchihewa, MW, Ranasinghe, USB, Weil, GJ. Reassessment of Areas with Persistent Lymphatic Filariasis Nine Years after Cessation of Mass Drug Administration in Sri Lanka. PLoS Neg Trop Dis. 2017;11(10):e0006066.

35. Joseph H, Maiava, F, Naseri, T, Silva, U, Lammie, P, Melrose, W. Epidemiological Assessment of Continuing Transmission of Lymphatic Filariasis in Samoa. Ann Trop Med Parasitol. 2011;105(8):567–78.

36. Weil GJ, Ramzy, RM. Diagnostic Tools for Filariasis Elimination Programs. Trends Parasitol. 2007;23(2):78–82.

37. Weil GJ, Curtis, KC, Fakoli, L, Fischer, K, Gankpala, L, Lammie, PJ, Majewski, AC, Pelletreau, S, Won, KY, Bolay, FK, Fischer, PU. Laboratory and Field Evaluation of a New Rapid Test for Detecting Wuchereria Bancrofti Antigen in Human Blood. Am J Trop Med Hyg. 2013;89(1):11–5.

38. Kubofcik J, Fink, DL, Nutman, TB. Identification of Wb123 as an Early and Specific Marker of Wuchereria Bancrofti Infection. PLoS Negl Trop Dis. 2012;6(12):e1930.

39. Steel C, Kubofcik, J, Ottesen, EA, Nutman, TB. Antibody to the Filarial Antigen Wb123 Reflects Reduced Transmission and Decreased Exposure in Children Born Following Single Mass Drug Administration (Mda). PLoS Negl Trop Dis 2012;6(12):e1940.

40. Weil GJ, Curtis, KC, Fischer, PU, Won, KY, Lammie, PJ, Joseph, H, Melrose, WD, Brattig, NW. A Multicenter Evaluation of a New Antibody Test Kit for Lymphatic Filariasis Employing Recombinant Brugia Malayi Antigen Bm-14. Acta Trop. 2011;120 Suppl 1:S19–22.

41. Gass K, Beau de Rochars, MV, Boakye, D, Bradley, M, Fischer, PU, Gyapong, J, Itoh, M, Ituaso-Conway, N, Joseph, H, Kyelem, D, Laney, SJ, Legrand, AM, Liyanage, TS, Melrose, W, Mohammed, K, Pilotte, N, Ottesen, EA, Plichart, C, Ramaiah, K, Rao, RU, Talbot, J, Weil, GJ, Williams, SA, Won, KY, Lammie, P. A Multicenter Evaluation of Diagnostic Tools to Define Endpoints for Programs to Eliminate Bancroftian Filariasis. PLoS Negl Trop Dis. 2012;6(1):e1479.

42. Dissanayake S, Xu, M, Nkenfou, C, Piessens, WF. Molecular Cloning and Serological Characterization of a Brugia Malayi Pepsin Inhibitor Homolog. Mol Biochem Parasitol. 1993;62(1):143–6.

43. Rao RU, Samarasekera, SD, Nagodavithana, KC, Punchihewa, MW, Ranasinghe, USB, Weil, GJ. Systematic Sampling of Adults as a Sensitive Means of Detecting Persistence of Lymphatic Filariasis Following Mass Drug Administration in Sri Lanka. PLoS Negl Trop Dis. 2019;13(4):e0007365.

44. Huppatz C, Durrheim, D, Lammie, P, Kelly, P, Melrose, W. Eliminating Lymphatic Filariasis--the Surveillance Challenge. Trop Med Int Health. 2008;13(3):292–4.

45. Ichimori K, Tupuimalagi-Toelupe, P, Toeaso Iosia, V. Graves, PM. Wuchereria Bancrofti Filariasis Control in Samoa before PacELF (Pacific Programme to Eliminate Lymphatic Filariasis). Trop Med Health. 2007;35(3):261–9.

